# The Role of Climate Change in the Expansion of Dengue

**DOI:** 10.1101/2025.10.06.25337235

**Authors:** Rafael de Abreu, Iago Perez Fernandez, Swapnil Mishra, Bernardo Gutierrez, Rhys P.D. Inward, Cathal Mills, Eduardo Lopez Ortiz, Leonardo S. Bastos, Laís Picinini Freitas, Luiz Max Carvalho, Seth Flaxman, Samir Bhatt, Samuel V. Scarpino, Flávio C. Coelho, Robert C. Reiner, Prathyush Sambaturu, Houriiyah Tegally, Simon Cauchemez, Oswaldo Gonçalves Cruz, César V. Munayco, José Alberto Díaz-Quiñonez, Dann Mitchell, Fraser Lott, Ben Lambert, William M. de Souza, Francesca Dominici, Oliver G. Pybus, Cláudia Torres Codeço, Marcia C. Castro, Moritz U.G. Kraemer, Sarah Sparrow

## Abstract

Climate change-related weather and extreme events are increasing in intensity and frequency(Nath and Palmer 2026), affecting infectious disease transmission globally(Mora et al. 2022). Dengue, a climate-sensitive vector-borne disease, to which over half the world’s population is at risk of infection, has expanded its geographical range over recent decades(Lim et al. 2025; Bhatt et al. 2013). The 2023/24 season marked the largest ever dengue outbreak year in the Americas, coinciding with the hottest year on record there(“Instituto Nacional de Meteorologia - INMET,” n.d.). Here, we use statistical models to investigate the Brazil 2023/24 dengue season and attribute how anthropogenic climate change impacted it. We analyze >20 years of dengue data across >5000 municipalities and find that observed temperature anomalies in 2023/24 increased dengue incidence in Brazil by 34% (95% CI: 22, 43%) accounting for population immunity. There was spatial heterogeneity across the country where in municipalities of southern Brazil temperatures were pushed into optimal thermal conditions for dengue transmission. For the best fitting model we found that in northern Brazil, 2023/24 temperatures became too high for effective transmission, resulting in lower dengue incidence compared to a counterfactual scenario without anthropogenic climate change, though with larger uncertainty. We test the generalizability of our model to high altitude areas in Mexico, where dengue has been expanding. Our work empirically demonstrates how a climate-change-related temperature anomaly led to the range expansion and growth of dengue across variable ecological and socio-economic settings, with implications for preparedness, adaptation, mitigation, and resilience planning.

## 1. Introduction

Climate change is driving changes in human health and will disproportionately affect populations in low- and middle-income countries(Dzau et al. 2025; Kristie L. Ebi et al. 2025; Smiley et al. 2022). Dengue virus (DENV), transmitted by *Aedes aegypti* and *Ae. albopictus* mosquitoes, has expanded its range in recent decades and it is estimated that more than half of the global population now lives in areas at risk of infection(Lim et al. 2025). The range and intensity of dengue outbreaks are expected to continue increasing(Intergovernmental Panel on Climate Change (IPCC) 2023) due to a set of co-occurring processes that include climate warming, climatic variability leading to more intense and frequent extreme events (such as longer warm and wet seasons, cyclones), repeated exposures to different DENV serotypes, increasing urbanization, and human mobility(Harish et al. 2024; Harris et al. 2024). In addition, climate patterns (e.g., *El Niño-Southern Oscillation*) and extreme events can alter the ecology and dynamics of DENV transmission, synchronizing epidemics across larger regions and/or causing severe outbreaks(Quandelacy et al. 2025; Dostal et al. 2022; Cazelles et al. 2023; van Panhuis et al. 2015; Mills et al. 2025). The focus of research to date linking weather and climate to dengue transmission has largely focused on how gradual changes in temperatures impact the global distribution of areas suitable for dengue transmission(Messina et al. 2019), and on potential changes in future dengue burden(Heath et al. 2025).

The 2023/24 dengue season in the Americas was the largest dengue epidemic on record there (>12 million cases and 7,000 deaths), with some countries and regions reporting substantial and unanticipated geographical expansion of dengue. In southern Brazil, the states of Paraná, Santa Catarina, and Rio Grande do Sul all declared health emergencies; the number of reported deaths in Rio Grande do Sul was larger than for all previous years combined(DATASUS 2025; Quandelacy et al. 2025; Codeco et al. 2022). The Pan American Health Organization(PAHO 2024) has suggested that this surge in cases was linked to Brazil experiencing its highest ever reported temperatures in 2023/24, 0.73°C above the historical average (1991 - 2020)(“Instituto Nacional de Meteorologia - INMET,” n.d., World Meteorological Organization 2025). However, the extent to which the 2023/24 temperature anomalies – and concurrent dengue epidemics – can be attributed to human-induced climate change remains unclear. Previous efforts to attribute human health outcomes to climate change(K. L. Ebi et al. 2025) have focused primarily on the direct effects of heatwaves on heat-related mortality with fewer studies on infectious diseases(Mitchell et al. 2016; Vicedo-Cabrera et al. 2023; Stuart-Smith et al. 2025; Barnes et al. 2025; Vicedo-Cabrera et al. 2021; Janoš et al. 2025; Carlson et al. 2025).

We seek to address this gap by analyzing >20 years of high-resolution, municipality-level time series dengue case data in Brazil, paired with climate and socioeconomic data. Our panel regression modelling framework accounts for local variation in climate seasonality, population immunity (estimated with force of infection models, seroprevalence data, and age-stratified case data), socio-economic (e.g., access to water and sanitation), and demographic characteristics. In a series of analyses we aim to (i) isolate the impact of climate-related drivers (i.e., temperature and precipitation) on dengue, (ii) evaluate the effects of the 2023/24 weather anomaly on dengue across regions, and (iii) use a state-of-the art climate model to quantify the relative contribution of anthropogenic climate-change to the record dengue incidence compared to a counterfactual scenario(Ciavarella et al. 2018).

## 2. Materials and Methods

### 2.1 Climate Data

In this study, we used monthly mean temperature from the ERA5 reanalysis (Hersbach et al. 2020), which provides an accurate representation of the past state of the atmosphere by combining past observations with weather prediction models. ERA5 has a spatial resolution of approximately 27 km and is available from 1940 onwards. For accumulated precipitation we used the Climate Hazards Group InfraRed Precipitation with Station (CHIRPS), which is available from 1981 onwards and has a spatial resolution of 5 km (Funk et al. 2015). We extracted data for each municipality over the study period and used these data to fit the model. If the municipality’s area was smaller than the model resolution, we used the value from the nearest grid point to the municipality centroid; otherwise, we calculated the average of the grid cells within the municipality.

To detect the contribution of anthropogenic climate change on the dengue epidemic, we used the UK Met Office HadGEM3-A model, which is run on an N216 grid (approximately 60 km in the midlatitudes). This model includes two scenarios: NAT, where only natural forcings are included—i.e., variability in solar irradiance at the top of the atmosphere and aerosols from volcanic eruptions—and ACT, where both natural and anthropogenic forcings are included—such as the effects of greenhouse gases, aerosols from industrial processes, and land use changes (Ciavarella et al. 2018). The ACT simulation is forced using observed sea surface temperature (SST) and sea ice extent from HadISST (N. A. Rayner, D. E. Parker, E. B. Horton, C. K. Folland, L. V. Alexander, D. P. Rowell, E. C. Kent, A. Kaplan 2003), while NAT has anthropogenic influence removed from SST and sea ice patterns based on delta derived from the Coupled Model Intercomparison Project Phase 5 (CMIP5) multi-model mean, as described in(Pall et al. 2011) and(Christidis et al. 2013). From this setup, 525 ensemble members are generated based on stochastic physics perturbations (Ciavarella et al. 2018), for each scenario for the year 2024, and are used to determine the differences between the two scenarios. This large number of ensemble members allows for better sampling of extreme events than what is currently available with coupled models(Ciavarella et al. 2018). We also use temperature and precipitation from HadGEM3-A historical runs, which span the period between 1960 to 2013 and have 15 ensemble members available. *El Niño* events alter regional climate patterns in the Americas and consequently dengue incidence. During 2023/24, a strong *El Niño* event contributed to reduced rainfall in the North and Northeast of Brazil, increased precipitation in the South, and widespread temperature rise across much of Brazil and South America (Cai et al. 2020). We explicitly consider the impact of *El Niño* including observed SST as a boundary condition in the HadGEM3-A model which ensures that the climate model reproduces the same *El Niño* phase as observed. By doing so we can isolate the impact of human-induced climate change on top of *El Niño*.

To ensure that our results are not systematically biased due to the HadGEM3-A model’s error, we applied a bias correction using ERA5 and CHIRPS data. For temperature, we employed the following equation:

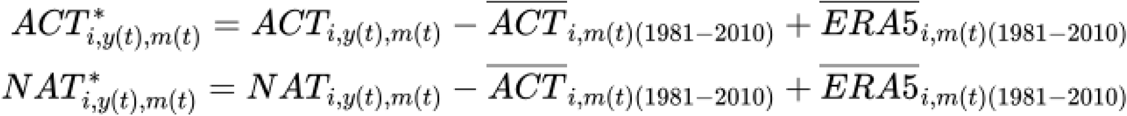

Where *i* represents the municipality, *t is the timestep, m* the month, and *y* the year. In this way, we subtract the monthly mean of the model over the 1981–2010 period and add the corresponding ERA5 monthly mean. For accumulated precipitation we divide by the climatology:

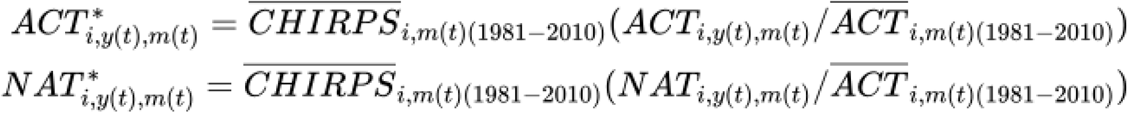

### 2.2 Dengue Case data

Brazil: Counts of probable dengue cases were obtained for each of the 5,570 municipalities in Brazil from Brazil’s Notifiable Diseases Information System (SINAN), available through the Health Information Department, DATASUS (https://datasus.saude.gov.br/informacoes-de-saude-tabnet/). The data are available from 2001 to the present, and are aggregated by month of the first symptom and city of residence. From DATASUS, we also extracted annual population estimates for each municipality. The data are further aggregated into the five distinct geo-political regions (North, Northeast, Southeast, South, and Centre-West) summing the number of cases in that region and dividing by the total population. We calculated the “center of mass” of dengue cases for each year by first computing the centroid of each municipality to obtain its *lon* and *lat* coordinates. We then calculated the weighted average of these coordinates using the number of cases as weights, according to the following equation:

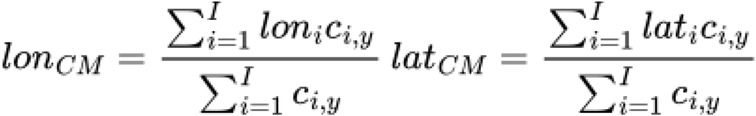

where *i* denotes the municipality, *y* is the year, and *c*_i,y_ is the number of cases in that particular year.

Mexico epidemiological data: For Mexico, we curated municipality-level data on confirmed dengue cases between 2020-24, defined as those identified through PCR or immunological diagnostic tests. These data are made publicly available by the Secretaría de Salud (Health Secretariat of Mexico) through its open-access epidemiological surveillance platform(de Salud, n.d.).

### 2.3 Socioeconomic, genomic, and immunological data and models

In addition to the climatic variables, we included other time-varying covariates in the model to assess the effects of socioeconomic conditions, genotype and serotype replacement and population immunity. The socioeconomic variables considered were the annual Gross Domestic Product (GDP) estimated for each municipality(Kummu et al. 2025), the percentage of urban area derived from MapBiomas(Souza et al. 2020), and the birth rate obtained from DATASUS. Furthermore, we fitted alternative model specifications including an indicator variable representing shifts in the dominant dengue serotype. This variable was coded as 1 (serotype shift) for the years 2008, 2012, 2013, 2019, 2020, and 2024, and 0 otherwise (Fig. S7). We also included annual lagged dengue case counts (from one to three years) for each municipality (Fig. S21).

To account for potential reporting biases across regions on reported cases we estimated the regional level annual population immunity using an age-stratified FOI model fitted to seroprevalence data updated in 12/2025 (Table S3). We used an extensive line list of dengue case data between 2002-24 sourced from the *Sistema de Informação de Agravos de Notificação* (SINAN)(“Portal de Dados Abertos Do SUS,” n.d.). As dengue is a mandatory notifiable disease in Brazil, this dataset includes laboratory-confirmed and clinical epidemiological cases reported to the Brazilian Ministry of Health through Sistema Único de Saúde. In short the model accounts for the proportion of severe dengue cases relative to total dengue cases, stratified by age, in order to estimate the proportion of individuals who had never been infected (Pr_0priorinf) and those who had been infected at least once (Pr_1priorinf), by region and year (Fig. S22). The inference model was based on a likelihood function of the annual force of infection (*λ*_*y,r*_) and probabilities of reporting dengue fever (*ϱ*_*F*_) or severe dengue (*ϱ*_*S*_), given data on annual reported cases of dengue fever (*F*_*y,r,a*_) and severe dengue (*S*_*y,r,a*_) stratified by year (*y*), region (*r*), and age (*a*). This likelihood function accounts for the population age distribution, as well as the probabilities that an individual’s first, second, or post-secondary dengue virus infection will result in dengue fever or severe dengue. These parameters are used to calculate expectations of each *F*_*y,r,a*_ and *S*_*y,r,a*_, with the data assumed to be independent Poisson random variables with those expectations. We obtained maximum-likelihood estimates (MLEs) of the model’s parameters in R version 4.3.2 (2023-10-31). MLEs of *λ*_*y,r*_ and the population age distribution were used to calculate predicted values of the proportion of individuals in each region in each year with no prior dengue virus exposures. Further details of the estimation procedure for these parameters are described by Brito et al.(Brito et al. 2021), with the estimates performed here simply being an update with additional years of data (2020-24) and seroprevalence studies included.

### 2.4 Statistical Model

To account for the contribution of known climatic drivers of dengue cases, temperature and precipitation, we adapted a panel regression model for monthly dengue cases proposed by(Childs et al. 2025) and commonly used in econometric analyses (Hsiang 2016). The Poisson Generalized Linear Model (GLM), as described in Equation 3 (Extended Data Table 1), is fitted using the **fixest** package in R (https://lrberge.github.io/fixest/). The model assumes that the number of cases in each municipality *i*, for each month *t* follows a Poisson distribution, where the expected value is a function of: temperature (*T*), represented by a third degree B-Spline with *k* = 4 degrees of freedom, with one knot at the median and boundary knots at the minimum and maximum, including lags 1 to 5 months; precipitation (*P*), represented by a linear term also including lags 1 to 5 months; an ɑ term that captures fixed effects for each municipality, thus accounting for places with generally higher case counts; δ, which represents fixed seasonal effects for each month *m(t)* of the year and for each considered region *r*; and *γ*, which represents fixed effects of interannual variability for year *y(t)* within each region. We also ran models with additional covariates as described in the previous section. We chose to separate the δ and *γ* terms by region (North, Northeast, Centre-West, Southeast, South) due to the large spatial extent of Brazil, where temperature and precipitation patterns vary significantly across regions. We used B-Spline with lags 1 to 5 instead of the third degree polynomial with lags from 1 to 3 as these are more flexible and provide better model performance (File S1). We also included a population offset in order to account for differences in population sizes; we standardized the offset by 100,000 so we model the rate by 100,000. To estimate the uncertainty associated with the model, we ran a spatial block bootstrap of 1,000 samples based on the states of Brazil.

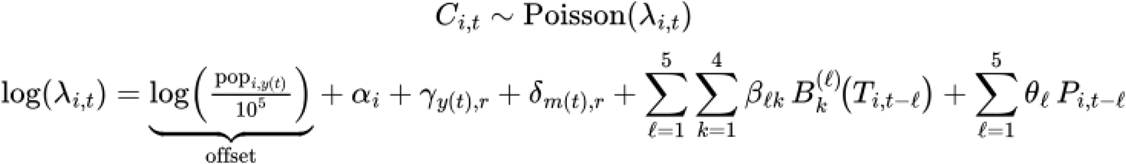

To estimate changes in dengue incidence for each region, we fit the model using ERA5 data and then use the estimated parameters to project cases for each of the 525 members of the HadGEM ensemble, under both ACT and NAT scenarios, at the municipal level. Subsequently, the number of cases was aggregated by region, summing across municipalities and dividing by the total population. For the values estimated using temperatures from 2005–2013 (Fig. 3; CLIM (2005–2013)), we used data from the 15 members of the historical runs while keeping the year term *γ* fixed at 2024, in order to estimate what the number of cases would be if conditions similar to 2024 were maintained, but with the temperatures from 2005-13. Our fixed effects specification controls for time-invariant unobserved confounders at the municipality level and potential temporal shocks. There are potential violations to the strict exogeneity assumption that include time-varying omitted variables such as vector control interventions and changes in health care utilization. However, because of the large sample, n = 5,570 municipalities and 20 years of data, we believe that our model is robust to detect the weather-dengue associations.

## 3. Results

### 3.1 Geographical expansion of dengue in 2023/2024 in Brazil

Annual incidence of dengue by municipality in 2023/24 (Fig. 1a; materials & methods) was concentrated in Brazil’s Southeast region, with ∼90% of municipalities there reporting monthly incidence rates >300 cases per 100,000 inhabitants (the threshold commonly used to define municipalities with high incidence(Lowe et al. 2014); Fig. 1b). High incidence was reported also in the Centre-West and South regions, with yearly incidence exceeding 4000 cases per 100,000. Together, these three regions account for >70% of the total population of Brazil and for the majority of domestic and international air travel. In contrast, the numbers of cases in the North and Northeast regions were lower, with only 30-40% of municipalities reporting >300 cases per 100,000 inhabitants (Fig. 1) and incidence similar to most previous years in the study period (Fig. 1d, e).

**Fig 1.**
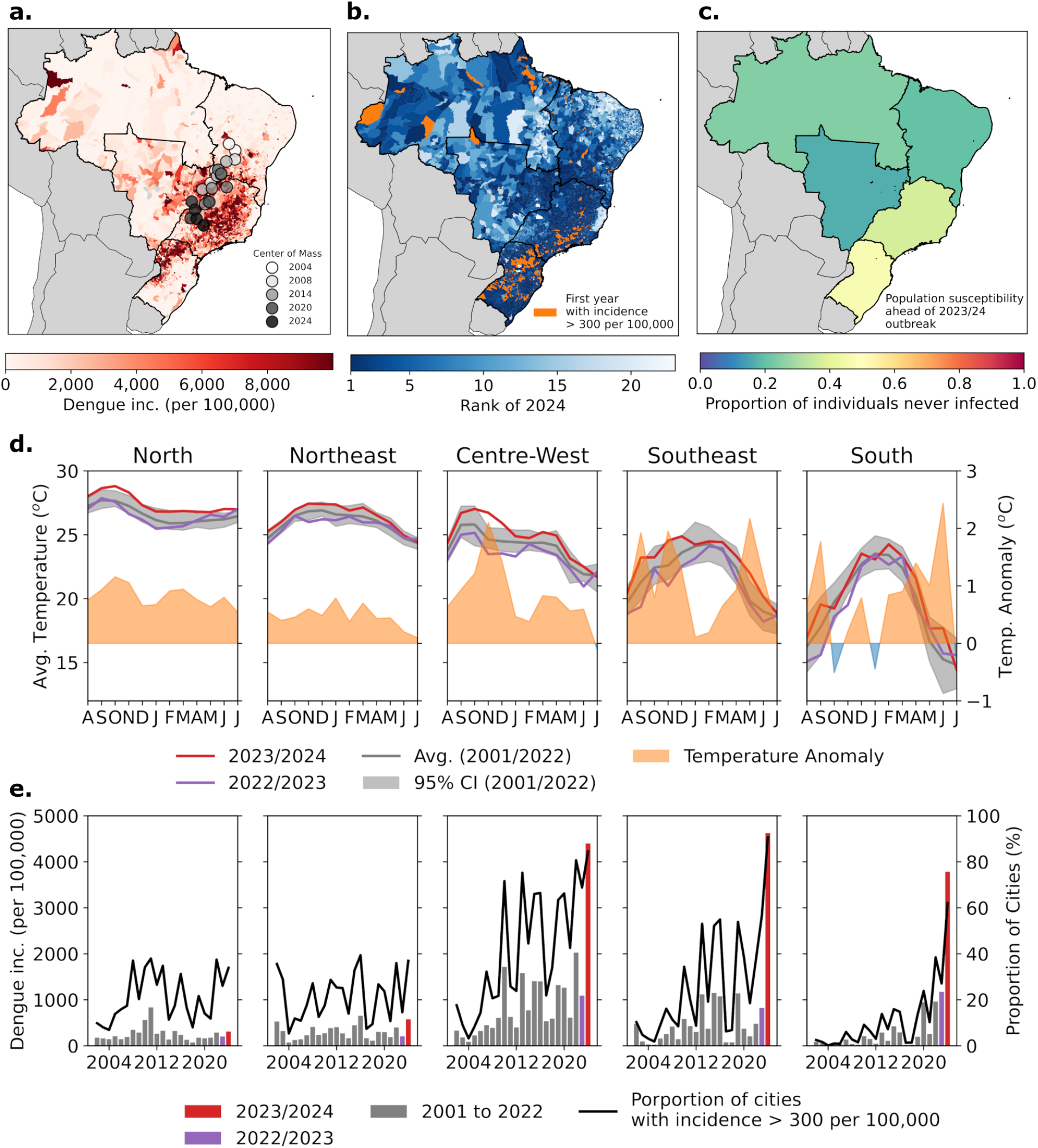
Twenty-year trends in dengue epidemiology and climate across Brazil. (a) Map of dengue incidence (per 100,000 inhabitants) between August 2023 and July 2024 per municipality. The black lines divide the country into five regions: South, Southeast, Centre-West, Northeast and North. The greyscale-shaded circles show the center of mass of dengue incidence per year, calculated as the average of the municipality coordinates weighted by dengue incidence. (b) Rank of dengue incidence per municipality in 2023/24 within study period, from 2004-24. Rank = 1 means that, in that municipality, 2023/24 was the year with the highest number of cases. Municipalities in orange are those that, in 2023/24, recorded for the first time >300 dengue cases per 100,000 in a year. (c) Estimated proportion of individuals never infected in ahead of 2023/24 season for each region (d) Population-weighted average temperatures for each Brazilian region, from August 2023 to July 2024 (ERA5 data; left hand axis). The red line shows values for 2023/24, and the purple line shows those for 2022/23. The black line shows the average across 2001-22 and the grey shading represents the 95% range of the interannual variability between 2001-22. The right-hand axis shows the temperature anomaly for 2023/24 compared to the average for 2001-22. Orange shading depicts higher temperatures in 2023/24 than the average, whilst blue shading represents lower than average temperatures. (e) Bar chart depicting the twenty-year time series of annual dengue incidence (per 100,000 inhabitants; left hand axis). The black line shows the proportion of municipalities in each region with >300 cases per 100,000 that year (right hand axis).

The center of mass of dengue incidence (calculated as the average of the municipality coordinates weighted by their dengue incidence) has moved southward over the last 20 years (Fig. 1a, shaded circles), so that by 2023/24 it had moved to the border of São Paulo and Minas Gerais states. During the same time, the center of mass of the Brazilian population remained largely stable (Fig. S1) despite an increase in the population of Brazil from ∼180M in 2004 to 210M in 2024.

Municipalities strongly affected by the 2023/24 DENV epidemic were located primarily in the Centre-West, Southeast and South regions of Brazil (Fig. 1a, b) with the South region experiencing their first major epidemic and the Centre-West and Southeast experiencing the largest ever. Population susceptibility might play a role in explaining variation in attack rate: we therefore reconstructed from high-resolution individual level and seroprevalence data the annual force of infection (FOI) and population susceptibility to dengue across regions in Brazil (materials & methods, Fig. 1c). We found that population susceptibility prior to 2023/24 was high in the South and Southeast (47 and 37 percent of the population were estimated not to have had any prior exposure to dengue, respectively) and comparable between the Centre-West, North and Northeast (16%, 24%, 20%, respectively; Fig. 1c). Our results are consistent with pre-2020 variation in population immunity(Brito et al. 2021) and track seroprevalence estimates (Extended Data Figs. 2-6). This makes the 2023/24 season an ideal scenario to test the impact of weather (temperature and precipitation) anomalies on dengue (i.e., regions with high incidence in 2023/24 had sufficient population susceptibility over the past decade and locations with comparable susceptibility had differences in attack rates). DENV genomic surveillance shows the invasion of DENV-3 genotype II (DENV-3II) during 2023, persisting at low prevalence through 2024(Pereira et al. 2025) and rising in frequency only during 2025. DENV-2 genotype II (DENV-2II) increased in relative abundance in Brazil from 2023 to 2024 affecting all regions simultaneously (from 35 to 50%), co-circulating with the already established DENV-1V (Fig. S7 & 8).

Using ERA5 climate data we observe above-average temperatures starting in August 2023 and continuing throughout the 2024 dengue season (typically December to June; Fig 1d; materials & methods) across all regions (Churakov et al. 2019). In the Southeast and Centre-West regions temperature anomalies reached +2°C in November 2023, compared to the average for 2001-22 (Fig. 1d). In the South region, temperature anomalies were less extreme at the end of 2023, but were consistently above +1°C at the beginning of 2024.

### 3.2 Reconstructing dengue transmission dynamics in Brazil

We estimated how climate, population susceptibility, and socio-economic factors impacted dengue in Brazil during 2023/24 using a fixed effects Poisson panel regression model (materials & methods; Fig. S9), in which we assume non-linear relationships between temperature and precipitation on dengue, fixed effects at the municipality level, and add additional annual population susceptibility from an age-stratified FOI model (materials & methods) and socio-economic factors (from(“IPS Brasil,” n.d., “PGI - Plataforma Geográfica Interativa,” n.d.)). The best fitting model reconstructing 20 years of dengue incidence across Brazil included lagged weather variables (monthly mean temperature and total precipitation), cumulative number of cases in previous year and reconstructed population immunity from seroprevalence data and age-stratified case data, serotype replacement information, and socioeconomic variables (urban percentage, GDP, and birth rate by municipality) as well as fixed effects for year and month (Fig. 2a, in sample adjusted Pseudo-R^2^ = 0.78; Squared Correlation = 0.68; lowest BIC (3.8987), File S1, Fig. S10). Our model generalizes well to municipalities and months that were not part of the model training (Extended Data Figs. 11 & 12). Correlations vary by region with the highest in the Centre-West, Southeast, and South (0.92, 0.98, and 0.96 respectively).

**Fig. 2.**
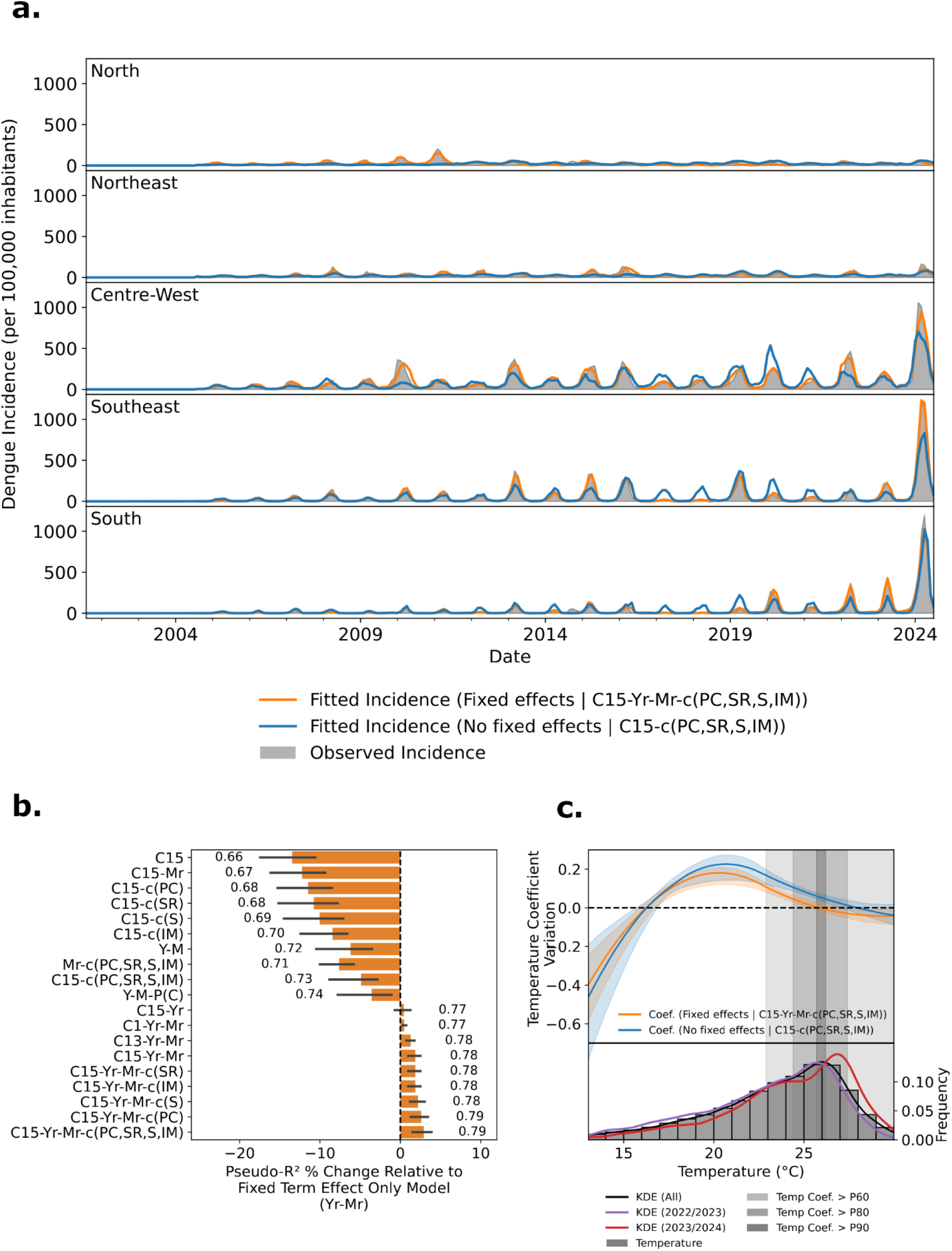
Modelling the relationship between temperature and dengue in Brazil. (a) Time series of monthly dengue incidence for each region in Brazil per 100,000 inhabitants. Grey shading shows reported dengue incidence; the orange line is the estimated cases from our Poisson regression model that includes year, month regional fixed terms, monthly temperature and precipitation lagged by 1-5 months, cases in prior year (rolling cumulative), serotype replacement indicator variable, socioeconomic variables, and population susceptibility estimates (C15-Yr-Mr-c(PC,SR,S,IM)); the blue line shows a model that is equivalent to the above except for the exclusion of fixed year and month terms (C15-c(PC,SR,S,IM)); (b) Change in Pseudo-R^2^ from the model with only fixed effects (i.e., only month, year, and municipality terms, without climate variables) shown as percentage change. The error bars represent a spatial block bootstrap of 1,000 samples, sampling from the different states in Brazil; (c) top panel shows the temperature-response curve for dengue extracted from the fitted Poisson regression model relative to the model’s intercept plus fixed effects for model C15-Yr-Mr-c(PC,SR,S,IM) and C15-c(PC,SR,S,IM) as in panel (a). Above zero means that dengue incidence is increasing with changes in temperature and below zero means that dengue incidence is declining. The shading shows 95% confidence intervals calculated using 1,000 bootstrap samples. The vertical greyscale shaded regions show the optimum temperature regions for dengue virus transmission based on the fitted model (from dark to light they represent the 90th, 80th and 60th percentile of the temperature coefficient of variation); (c, bottom panel) Histogram and Kernel Density Estimate (KDE) of temperature across municipalities for the entire year. The purple line shows 2022/23 and the red line is 2023/24. Grey bars show averages from 2004-23. Model abbreviation: C1: climate variables (lag 1), C13: climate variables (lag 1-3), C15: climate variables (lag 1-5), P(C): 3rd degree polynomial (Climate lag 1–3), M: month fixed effect, Mr: month fixed effect per region, Y: year fixed effect, Yr: year fixed effect per region; c(…) stands for additional covariates like: PC: Cases from prior years (lag 1-3 years); SR: Serotype replacement indicator variable; S: Socioeconomic variables (births, GDP, and urban area); IM: Immunity.

We estimate, similar to others(Lowe et al. 2021), a strong non-linear relationship between lagged mean monthly temperature and dengue incidence with an increase in the effect of temperature on dengue incidence up to 26°C (with the largest marginal increase of dengue estimated around 21°C; values above zero mean higher values of temperature are associated with higher dengue relative to the baseline vs. values below zero are associated with lower dengue incidence) and a decrease above that temperature, albeit with a less strong decline than previous estimates from long-term historic and global data (Fig. 2c)(Childs et al. 2025). A version of the model without the inclusion of fixed effects was able to reconstruct well the observed patterns aside from the larger peaks and the temperature response curve remained similar to that with fixed effects (Fig. 2c). There is regional variation in temperature response curves when models were fitted independently due to differences in observed temperature regimes across the wide geographical regions in Brazil (Fig. S13). Regional models reveal variation in estimated peak temperature-dengue relationships; the North and Northeast (25.5C, 27C respectively) tend to be higher than in the Centre-West, South, and Southeast (24, 24.5, 25 respectively, Fig. S13). Annual variation in temperatures between 2002-24 shows predictable relationships in high incidence years with stronger variation when few cases were reported (Fig. S14).

We estimated that, during 2023/24, the temperature in some municipalities was too high for optimal transmission as estimated from our data but within range based on estimates from laboratory studies (Fig 2c, red line)(Mordecai et al. 2017). Estimates of the temperature-dengue relationship are robust to changes in the model configuration (e.g., fixed vs. random effects, Table S2, Fig. S15).

### 3.3 Variation in Brazilian dengue incidence that can be attributed to anthropogenic temperature change

We next sought to decompose dengue incidence in 2023/24 into the fraction attributable to human-induced climate change, versus natural variability in the climate system while accounting for variation across other known dengue drivers. We used the HadGEM3-A system, which implements climate simulations that can represent (i) the climate as it is, including anthropogenic forcings and local climate patterns (ACT), and (ii) how the climate would be without human-induced change (NAT). The HadGEM3 model was designed for climate attribution studies and is used to investigate the contribution of anthropogenic climate change to the frequency and intensity of climate extreme events(Christidis et al. 2013; Ciavarella et al. 2018; “Climate Change, El Niño and Infrastructure Failures behind Massive Floods in Southern Brazil – World Weather Attribution,” n.d.). The difference in temperature between the ACT and NAT scenarios can be viewed as the temperature change attributable to anthropogenic action (i.e., greenhouse gas and anthropogenic aerosol emissions, and land use changes)(Christidis et al. 2013; Ciavarella et al. 2018).

In addition to the different forcings, the model ensembles are initialized with fixed boundary conditions; specifically, observed sea surface temperature (SST) and sea ice coverage (SIC; HadISST1 dataset(N. A. Rayner, D. E. Parker, E. B. Horton, C. K. Folland, L. V. Alexander, D. P. Rowell, E. C. Kent, A. Kaplan 2003)). For NAT, the ensembles are initialized with the same boundary conditions as for ACT but with an estimate of the anthropogenic attribution on SST and SIC removed. Crucially, both scenarios are not biased by variation in the climate system such as *El Niño(Pirani et al. 2024; Araujo et al. 2025)*.

To validate the temperatures modelled using HadGEM3-A we compared them to those derived from ERA5 observational reanalysis; the two models align well during the 2023/24 study period (Fig. 3a). Importantly, we find that in the North and Northeast of Brazil the observational data and the model with anthropogenic forcing (ACT, orange) exhibit temperatures *above* the estimated thermal optimum for dengue transmission. In contrast, in the Southeast and South of Brazil, the model with anthropogenic forcing (ACT) has raised temperatures such that they fall within the optimal thermal regime for DENV transmission, whilst temperatures in the mode without anthropogenic forcing (NAT, blue) remain below the optimal range. The situation for the Centre-West region is intermediate between that observed for the abovementioned northern and southern regions. In summary, thermal conditions similar to 2023/24 were expected to occur every 6.84 (95% CI: 5.95, 8.19) years in a scenario with human-induced climate change (ACT), whereas the likelihood of such conditions is close to zero in the scenario without anthropogenic forcing (Fig. S16).

**Fig. 3.**
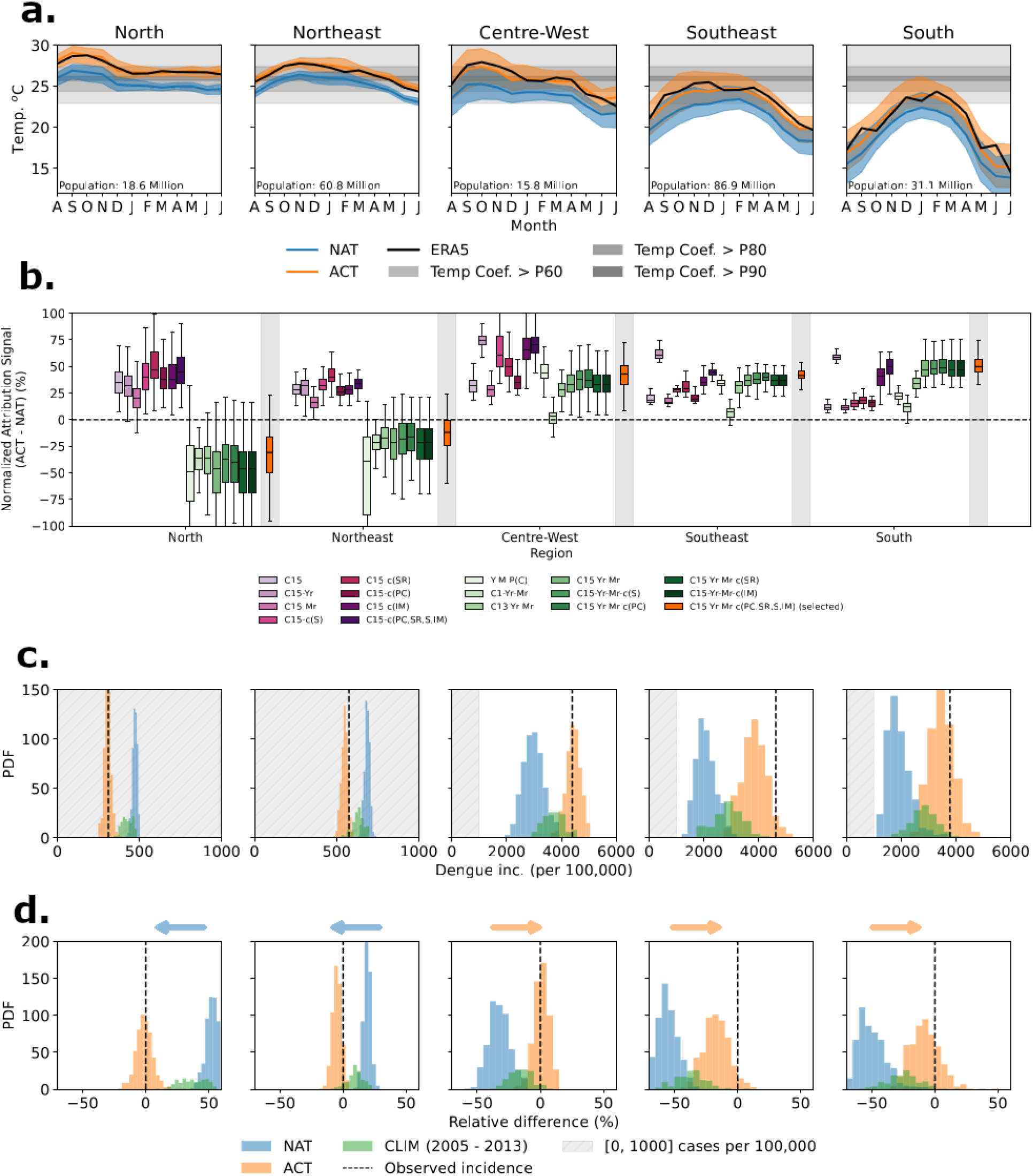
Quantification of the effect of anthropogenic climate change on dengue incidence. (a) Average monthly temperature between August 2023 and July 2024 for the different regions in Brazil, for the ACT (orange) and NAT (blue) models. The observation-based ERA5 temperatures are shown also (black). The horizontal greyscale shaded regions show the temperature regions for dengue virus transmission based on the fitted model (from dark to light they represent the 90th, 80th and 60th percentile of the temperature coefficient of variation; Fig. 2). (b) Percentage differences in estimated dengue cases using temperature values from the ACT and NAT scenarios. Each bar shows a different model specification, including the selected model in red (shaded background). Boxes show the interquartile range, center line the median and whiskers extend to the 95% CIs.; (c) For each Brazilian region, histograms show the distribution of inferred dengue incidence (cases per 100,000 inhabitants) across the 525 ACT and NAT simulations. The horizontal axis scale differs among panels; grey shaded region indicates the range 0-1000. We also estimated incidences using estimated temperatures from 2005–13 (CLIM0513; green). (d) Same as (b), but each histogram shows the difference between the dengue incidence inferred under the ACT (orange) or NAT (blue) models. Absolute differences were divided by observed incidence to obtain a relative difference metric. The arrows above each plot show whether dengue incidence increased or decreased in that region, compared to the scenario with no anthropogenic climate change (NAT).

Dengue incidence by region estimated from our model using the climate data from the ACT ensemble closely matches the observed cases (Fig. 3c). The construction of our panel regression model means that we can quantify how differences in temperature across the different climate scenarios impact dengue incidences among regions(Barcellos et al. 2024; Lowe et al. 2021; Mills and Donnelly 2024). By comparing the difference between the mean temperatures of the ACT and NAT distributions, we estimate an increase in dengue cases due to human-induced temperature change of 34.7 % (95 % CI: 22.3, 42.7 %) in Brazil, and of up to 50% in the Centre-West, Southeast, and South regions, with the largest increase recorded in the latter, averaging 49.8 % (95% CI: 36.1, 74.3) using our best best-fitting model (Fig. 3b). As noted above, this is likely due to temperatures there moving toward values more conducive to mosquito reproduction, virus replication, and transmission(Siraj et al. 2017). Furthermore, we estimate that the probability of an event of this magnitude occurring without anthropogenic induced climate change (i.e., the NAT scenario) is close to zero in the South, Southeast and Centre-West regions (Fig. 3c,d). This result is consistent across different model sensitivity tests for the Centre-West, Southeast, and South regions, showing an overall positive signal (Fig. 3b). The model considering temperatures lagged by one month shows the lowest attribution signal, likely due to that model’s reduced temperature sensitivity (Fig. S4a), whereas the model including only climate variables and fixed effects for the year shows the strongest signal in these regions.

In the North and Northeast regions our findings are more variable depending on the model chosen. In best fitting models with year and month fixed effects (green and red), we found that temperatures were too high for effective transmission and therefore a reduction of cases can be observed for the ACT scenario compared to the NAT scenario (average decrease of -31.0 % (95% CI: -81.8, 11.4%) in the North and -12.0 % (95% CI: -51.8, 15.4%) in the Northeast). The reason for the differences in estimates might be the high temperatures all year round in the northern regions though transmission peaking mostly in the southern hemisphere summer months (December - March). The higher population density in the southern regions, combined with the more favorable thermal conditions for transmission, therefore, increased the potential for severe dengue outbreaks such as the one observed in 2023/24.

In our analysis total monthly precipitation did not improve model fit (Fig. S17) which might be due to the timescale used in our analysis (monthly). We tested the signal of the anthropogenic effect using different specifications based on the C15-Yr-Mr model, that is, including fixed effects for year and month by region, a municipality fixed term, and climatic variables with lags from 1 to 5. Running the model using the average temperature between 2003 and 2013 with precipitation from 2023/2024, we observed that the difference between the two scenarios was almost zero. In contrast, keeping precipitation fixed between 2003 and 2013 but using temperatures from 2023/2024 yields results similar to those obtained when using 2023/2024 data for both variables. In addition, the estimates remain consistent when assuming a non-linear relationship.

### 3.4 Model comparison and validation

While other climate attribution studies have focused primarily on comparisons between contemporary and pre-industrial climate conditions, several factors beyond climate are likely to differ between those times, including population structure, urbanization, and international travel. In order to model a more conservative scenario, we compared our results from ACT with temperature estimates from the historical runs of HadGEM3-A for the years 2005–13 (CLIM0513), a period prior to the large Zika outbreak in Brazil but with substantial impact of anthropogenic climate-change(Faria et al. 2017, 2016; Brito et al. 2021). We chose this period because ZIKV infection has been shown to enhance subsequent severe dengue disease for some DENV serotypes while protecting against other serotypes(Katzelnick, Bos, et al. 2020; Katzelnick, Narvaez, et al. 2020). In this analysis we still estimate an increase of ∼20% of dengue cases in 2023/24 compared to the reference period in the Centre-West, Southeast, and South regions.

As an additional validation, we note that areas in the Americas have experienced recent dengue expansion; in Mexico the proportion of dengue-affected municipalities increased from 38% to 68% between 2022-24, with marked expansion into higher altitude areas(Mendoza-Cano et al. 2025). We obtained municipality level data from Mexico and tested our model’s generalizability to high altitude areas (>1000 meters) in that country. Our model accurately predicts increasing cases of dengue due to increases in temperature and reduced incidence in areas <1000 meters in 2023/24 (see Fig. S18).

## 4. Discussion

Our results show that climate change-related temperature anomalies can reshape the transmission dynamics of DENV. In Brazil, we show that they increased transmission in highly populated regions in the Centre-West, Southeast and South and decreased transmission in Northern regions. Newly affected areas might become hotspots of transmission and sources of further DENV geographic dissemination(Poongavanan et al. 2024); a record number of imported dengue cases have been reported in Europe in 2023/24(European Centre for Disease Prevention and Control 2024). Between 2004-24, Brazilian international and domestic air passenger flows tripled and quadrupled, respectively (0.4 to 1.2M international passengers; 2 to 8M domestic passengers). This could lead to greater risks of DENV importation and exportation (Fig. S19 & 20), especially as Southeast Brazil is where the majority of international (80%) and domestic (50%) passengers arrive and depart.

Our model indicates that DENV incidence peaks at approximately 26°C while in idealized conditions in the laboratory it has been suggested that R_0_ peaks at higher temperatures (estimated at 29°C for *Ae. aegypti(Athni et al. 2024)*). Our work contributes to the growing evidence of the impacts of climate on infectious disease transmission, which are also influenced by limited intervention options, changes in socio-economic conditions, and health-care disruption(Laydon et al. 2025). We identified a strong non-linear relationship between temperature and dengue which made fewer assumptions than previous studies using panel regression methods that typically assumed more constrained functional forms resulting in rapid decrease in transmission after the peak(Wagner et al. 2020). Recent studies have emphasized the cascading impact of climate extreme events on healthcare disruption(Symons et al. 2025; Rice et al. 2025), which might compound the documented direct impacts of temperature on health. Assessing the relative contributions of ecological and secondary (e.g. health-care disruption) related impacts will likely depend on the event and the economic and social context(Tsui et al. 2024).

The 2024 epidemic was estimated to cost >USD$5 billion, including healthcare costs and losses in production and services(Vancini et al. 2024) and underscores the social and economic relevance of climate change healthcare impacts in Brazil. Adaptation and mitigation measures are needed to reduce the impact of dengue across the country and globally(Camprubí-Ferrer et al. 2026). Measures to improve resilience include improvements in infrastructure (including constant water availability), ensuring sustainable urban growth(Gibb et al. 2023), education(MacCormack-Gelles et al. 2018), and pre-emptive and effective use of vector control(Padilha et al. 2025).

Rollout of the Qdenga Dengue vaccine by Takeda might reduce the number of infections in high-incidence regions by 20%(Cracknell Daniels et al. 2025) but there remain substantial challenges to large-scale rollouts(Scott et al. 2023), emphasizing the importance of continued investments in prevention, adaptation and mitigation(Ponmattam et al. 2025; Hadisoemarto and Castro 2013). The tetravalent Butantan Dengue vaccine (Butantan-DV) with a single-dose regime showed an efficacy of ∼80% against DENV-1 and DENV-2 in a phase 3 trial with a two-year follow-up, but detailed studies of seronegative individuals are still underway(Kallás et al. 2024). How DENV serotype co-circulation might have contributed to the increasing burden of dengue in Brazil remains unanswered; in this work we did not find indication that the large 2023/24 outbreak was linked to a sero- or genotype replacement event (Fig. S7 & 8). Additional data on temporal trends in population and geno- and serotype specific immunity at high spatial resolutions would improve our understanding of the impact of immunity on the spatial epidemiology of dengue(Mina et al. 2020).

In our analysis we were able to explicitly account for population immunity which was lower in the South and Southeast regions prior to the large epidemic in 2023/24. That means that there were sufficient possibilities in the past for epidemics to occur there had the climate conditions been favourable. Consequently, our results of lower transmission in the Northeast and North and higher transmission in the South, Southeast and Centre West in the 2023/24 season are best explained by changes in temperatures that allowed epidemics to grow in southern regions where a large number of populations were susceptible to infection. We also document that temperature-dengue relationships can be regionally specific (Fig. S13) but overall that the model generalises well to other settings.

Our work has several limitations. Dengue transmission and disease burden is driven by a complex interplay of factors including human behavior, urbanization, socio-economics, heterogeneity in vector control(Ferrari 2023), prior immunity, serotype/genotype circulation, and clinical vulnerability. For example, the North and Northeast regions, where we identified a decrease in cases due to climate change, are projected to experience more droughts in the future(Jeferson de Medeiros et al. 2022)(Jeferson de Medeiros et al. 2022), may lead to more water storage in open tanks, creating additional breeding sites in proximity to human hosts for *Aedes* mosquitoes(Rufasto Goche et al. 2025; Lowe et al. 2021). There have also been reports of rising prevalence of DENV-3 in Pernambuco in the Northeast in 2025 which might result in a wave of infection(Santos 2025). Other drivers of DENV transmission that we were unable to account for include time-varying healthcare testing capacity and seeking behavior and human mobility(Romeo-Aznar et al. 2022; Bomfim et al. 2020). However, we believe that many of these additional factors are already accounted for in the municipality fixed effects. There are uncertainties related to climate reanalysis and climate model data that might impact our estimated temperature-dengue relationship and why it differs from laboratory data (e.g. coarser resolution of climate data and local heat islands in cities). However, our results are consistent with previous work in the region(Harish et al. 2024) and our modelling framework and estimates provide evidence of similar patterns in other settings (Fig. S18). Future work might focus on the attribution of multiple climate extreme events to dengue outbreaks and to infectious diseases more generally(Quilcaille et al. 2025). While AI models are becoming better in anticipating weather extreme events(Meng et al. 2025), translating it into cascading risk for climate-sensitive diseases remains challenging. Mitigating and adapting to climate extreme events in regions nearing favorable conditions for dengue transmission will be essential for reducing the avoidable future burden of dengue.

## Data Availability

All data and code to reproduce the results are available from https://github.com/rafaelcabreu/dengue_attribution

## Acknowledgments

We thank the JASMIN team and Centre for Environmental Data Analysis (CEDA) for providing the computational infrastructure used for data processing. We want to thank T. Alex Perkins for helping out with estimating population immunity.

## Author contributions

Conceptualization: R.dA., S.S., M.U.G.K. Methodology: R.dA., I.P.F., S.F., S.M., L.S.B., C.T.C., L.M.C., S.B., S.V.S., F.C.C., R.C.R., F.D., M.C.C., D.M., S.S., M.U.G.K. Data curation: R.dA., M.U.G.K., E.L.O., I.P.F., C.M.E., B.G., R.P.D.I., J.A.D-Q., P.S. Investigation: R.dA., I.P.F., S.M., T.A.P., M.U.G.K. Visualization: R.dA., M.U.G.K., R.C.R. Funding acquisition: S.S., M.U.G.K. Project administration: R.dA., S.S., M.U.G.K. Supervision: S.S., M.U.G.K., M.C.C. Writing – original draft: R.dA., M.U.G.K., M.C.C. Writing – review & editing: I.P.F., S.M., B.G., R.P.D.I., E.L.O., L.P.F., O.G.C., C.M.E., L.S.B., C.T.C., L.M.C., S.B., S.V.S., H.T., F.C.C., R.C.R., O.G.P., F.D., M.C.C., D.M., S.S.

## Supplementary Materials

### Sensitivity analyses

The results for the fitted model are in File S2 and Fig. 2a, showing good agreement with the observation. Still, we conducted sensitivity tests to assess the robustness of the model results. The models used and their formulation are included in Table S1. We also tested whether the temperature-dengue coefficients are consistent with a model with random effects for month, year and municipality instead of fixed terms using a Bayesian model fitted with The Integrated Nested Laplace Approximation (INLA) package in R (https://www.r-inla.org/). In this case the random effects were assumed to be iid variables for municipality and year, and a cyclic random walk term for the month. We did not include spatial autocorrelation between municipalities due to computational complexities.

File S2 shows the fitted coefficients for each of these models as Incidence Rate Ratios (IRR) and summary statistics of the different models tested, highlighting in gray the model used in the main manuscript. We compared the inclusion of the climate terms related to the fixed effects only model (Yr-Mr). Compared to this model, we observe that the model C15-Yr-Mrhas a lower AIC and a higher Pseudo-R^2^. The best fitted model, chosen to do the analysis in the main text, is C15-Yr-Mr-c(PC,SR,S,IM), in which temperature with lags from 1 to 5 months was included, as well as additional covariates for prior cases, serotype replacement, socioeconomic indicators, and immunity estimate with a higher Pseudo-R2 and lower AIC and BIC. The model with a third degree polynomial, and no fixed effects for region, denoted as Y-M-P(C) had a higher AIC and lower Pseudo-R^2^ when compared with models with regional fixed terms, justifying the disaggregation of month and year terms by region due to the reduction in error.

We also evaluated the change in the temperature response using models that include the climate terms (Fig. S15a). We observed that the shape of the curve remains consistent across most sensitivity tests, with the peak incidence occurring between 26 and 27 degrees. Even when we fit the function using different datasets, such as those from Mexico, the estimated thermal optimum remains close to 27 degrees. In the case of precipitation (Fig. S15b), we observe in general positive values indicating that increased rainfall raises the number of cases; however, the uncertainty is high with the data from Mexico showing a negative relationship between rainfall and dengue (similar to observations in other work(Lowe et al. 2021)).

### 2.6 Comparison of attributable signal

In the case of the difference in regional incidence between the ACT and NAT scenarios presented in Fig. 3b, we calculate that by running a spatial block bootstrap of 100 samples based on the states of Brazil for each of the 525 ensemble members of the ACT and NAT scenarios, which gives the contribution of the climate variables to the model result. After that we calculate for each bootstrap sample the average contribution for ACT and NAT, by averaging across all ensembles, and then calculate the difference normalized by the observed incidence.

#### México validation study

We also included data from municipalities in Mexico in the model fitting to evaluate its performance. As shown in Fig. S18b, dengue incidence in 2024 was calculated for Mexico between January and December 2024, instead of August to July as in Brazil due to the seasonal differences between the Northern and Southern Hemispheres. Most cases in Mexico are concentrated in low-altitude coastal regions, while the continental interior, where altitudes are higher (Fig. S18a), records fewer cases. Similar to what was observed in the Centre-West, Southeast, and South regions of Brazil, the above 1000 m region exhibits ACT temperatures close to ERA5, indicating that the model accurately captures both temperature and seasonal cycles in these areas, with ACT being closer than NAT to the optimal temperature for dengue transmission (Fig. S18c), which suggests an increase in transmission. This results in an increase in the number of cases, which in the case of the above 1000 m region in Mexico is close to 50% and aligns with studies projecting an increase in dengue incidence in high-altitude areas of Mexico (Harish et al. 2024).

#### DENV genomic data

We downloaded all available Brazilian sequences and their associated metadata between 2004-01-01 and 2025-01-01 from GenBank(Benson et al. 2013) and GISAID (Shu and McCauley 2017) (data downloaded on 2025-08-20). Sequences missing essential metadata, such as collection date or location, were removed. As there is no linkage ID between GenBank and GISAID, to remove duplicates sequences, sequences from both datasets were screened for identity and, in the case of identical sequences (those with the same location, collection date, and sequence), only one such isolate was retained. Due to errors in serotype labelling within the GenBank metadata, we used Dengue Lineages(Hill et al. 2024) to identify the serotype, genotype, and sub-lineage of each sequence.

#### Passenger flight data and processing

We downloaded monthly passenger flight data from 01/2021 - 12/2024 from the official government flight data source, the Brazilian Agência Nacional de Aviação Civil (Anac) data portal(Agência Nacional de Aviação Civil (Anac), n.d.). Data were summarized for all flights arriving and departing from airports in Brazil and aggregated to the North, Northeast, Centre-West, Southeast, South regions. Data represent passengers on direct routes only, meaning that we do not know the origin locations of each of the incoming or outgoing passengers.

**Table S1.**
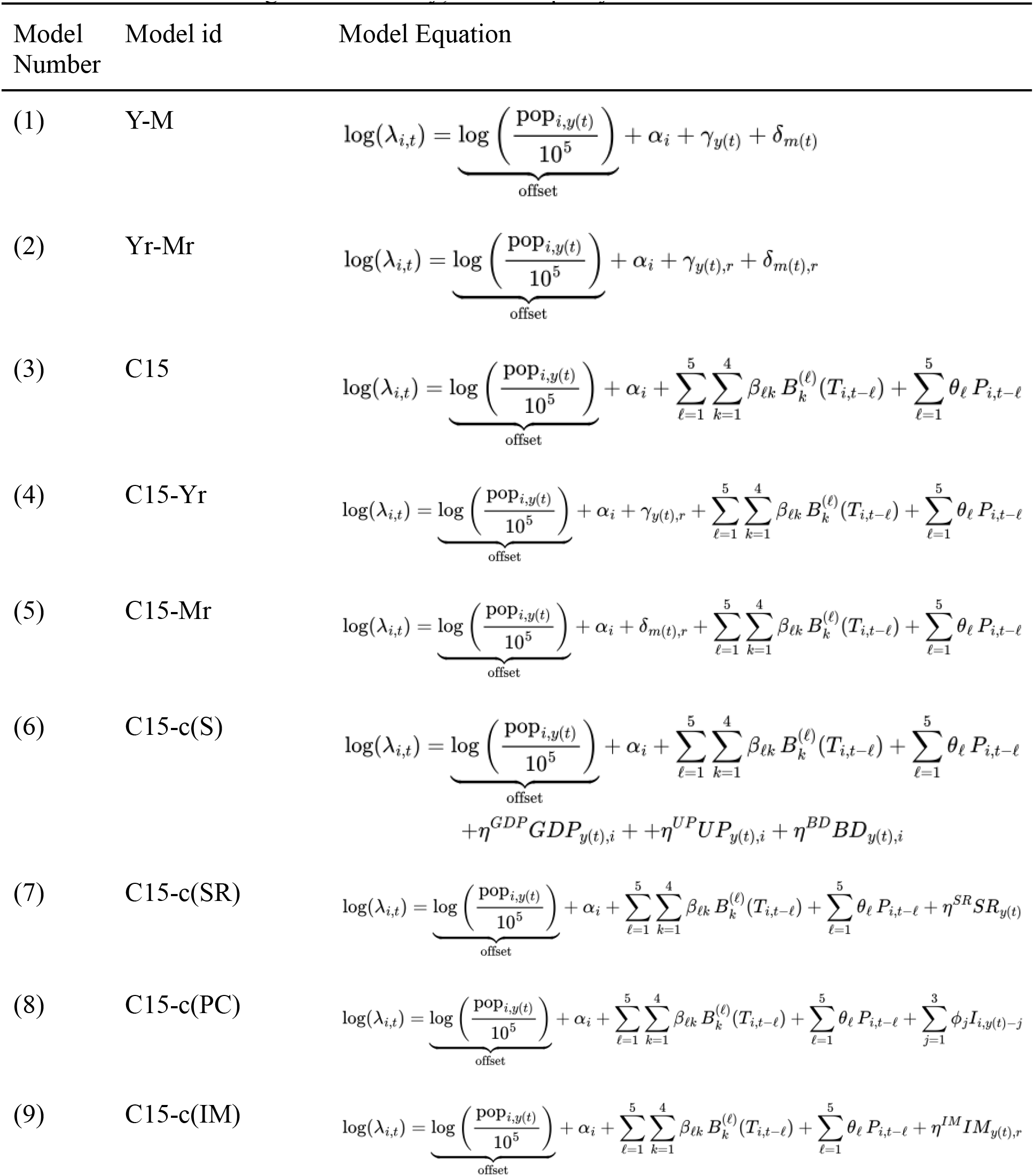
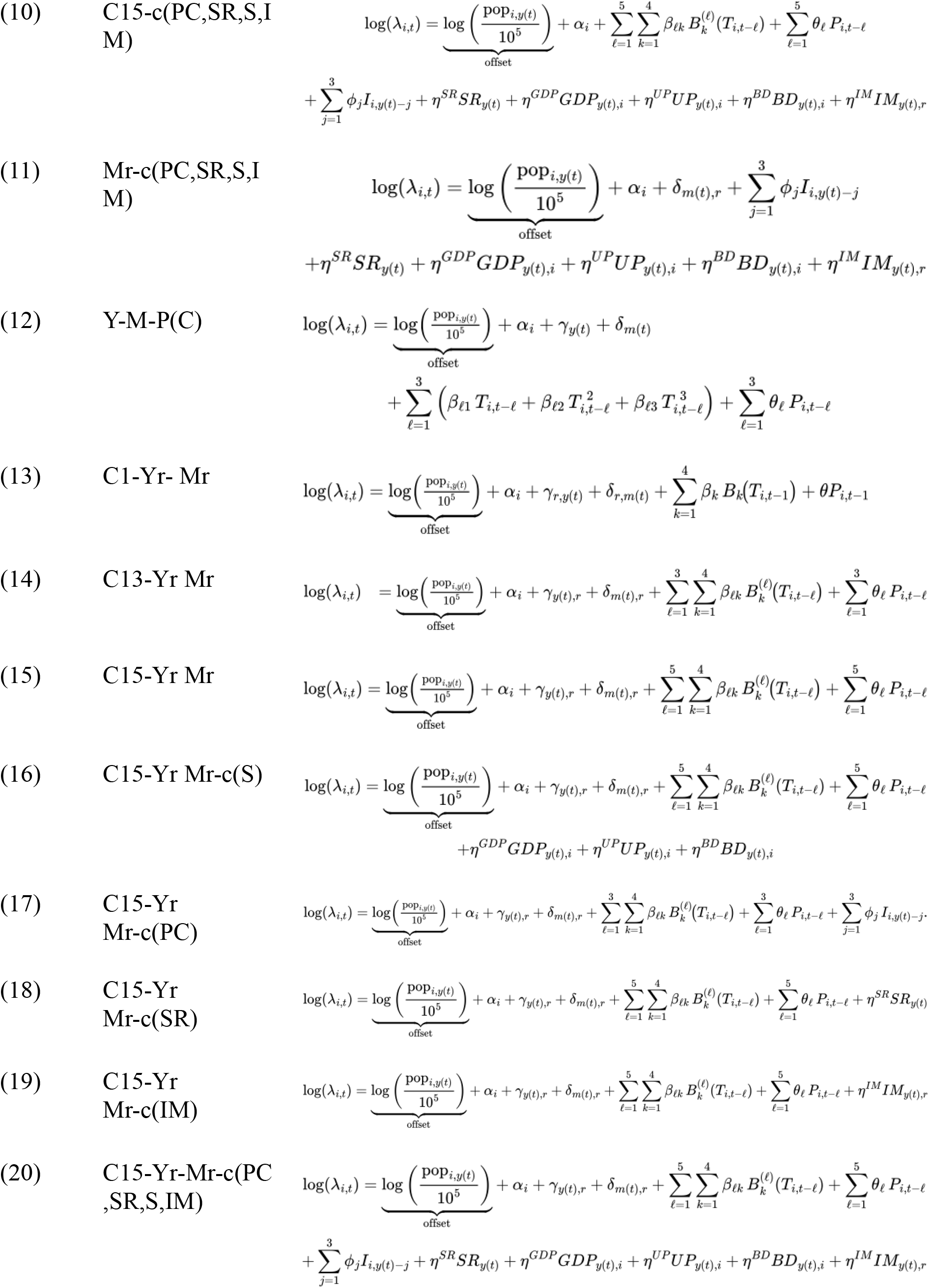
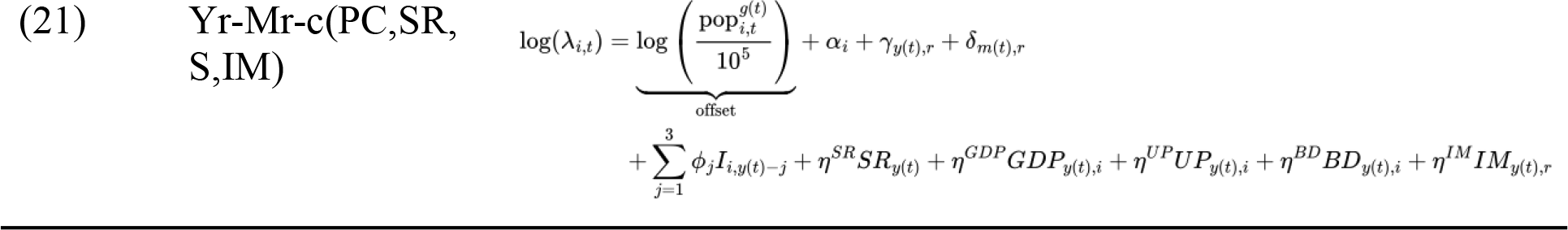
Model number, name used in the text and equation. The model abbreviations are: Y: year fixed effects year *γ(t)*, Yr: Region-specific year fixed effects *γ(t)*_*r*_, M: monthly fixed effects *δ(t)*, Mr: Region-specific monthly fixed effects *δ(t)*_*r*_, C1, C13, C15: Temperature terms (*T*) modeled using B-splines with lags and precipitation as a linear fit (*P*); P(C): Third degree polynomial for temperature and precipitation as a linear fit, c(·): Additional covariates included in the model: S: Indicator of socioeconomic variables (*GDP* gross domestic product per capita, *UP* urban percentage, *BD* birth rate); PC lagged annual cases *I*_*i,y(t)-j*_; SR: Serotype Replacement indicator variable IM: regional immunity; *α* municipality fixed effect.

**File S1**. Incidence Rate Ratios (IRR) of the variables for the fitted models in the sensitivity test and statistical significance (‘***’ p-value < 0.001 ‘**’ p-value < 0.01 ‘*’ p-value < 0.05). “BS” represents the b-splines for each of the lagged terms. Below is also the Akaike information criteria (AIC), Bayesian information criteria (BIC), Squared Correlation, and pseudo R2 for each model in our sensitivity analysis. The grey shading represents the selected model for the main text. Link for the table: https://docs.google.com/spreadsheets/d/1zJ-NdFpnFg1Yu6iP44ShtIsIVjhjMvZ4FeoGKwZtsHk/edit?usp=sharing

**Table S2.**
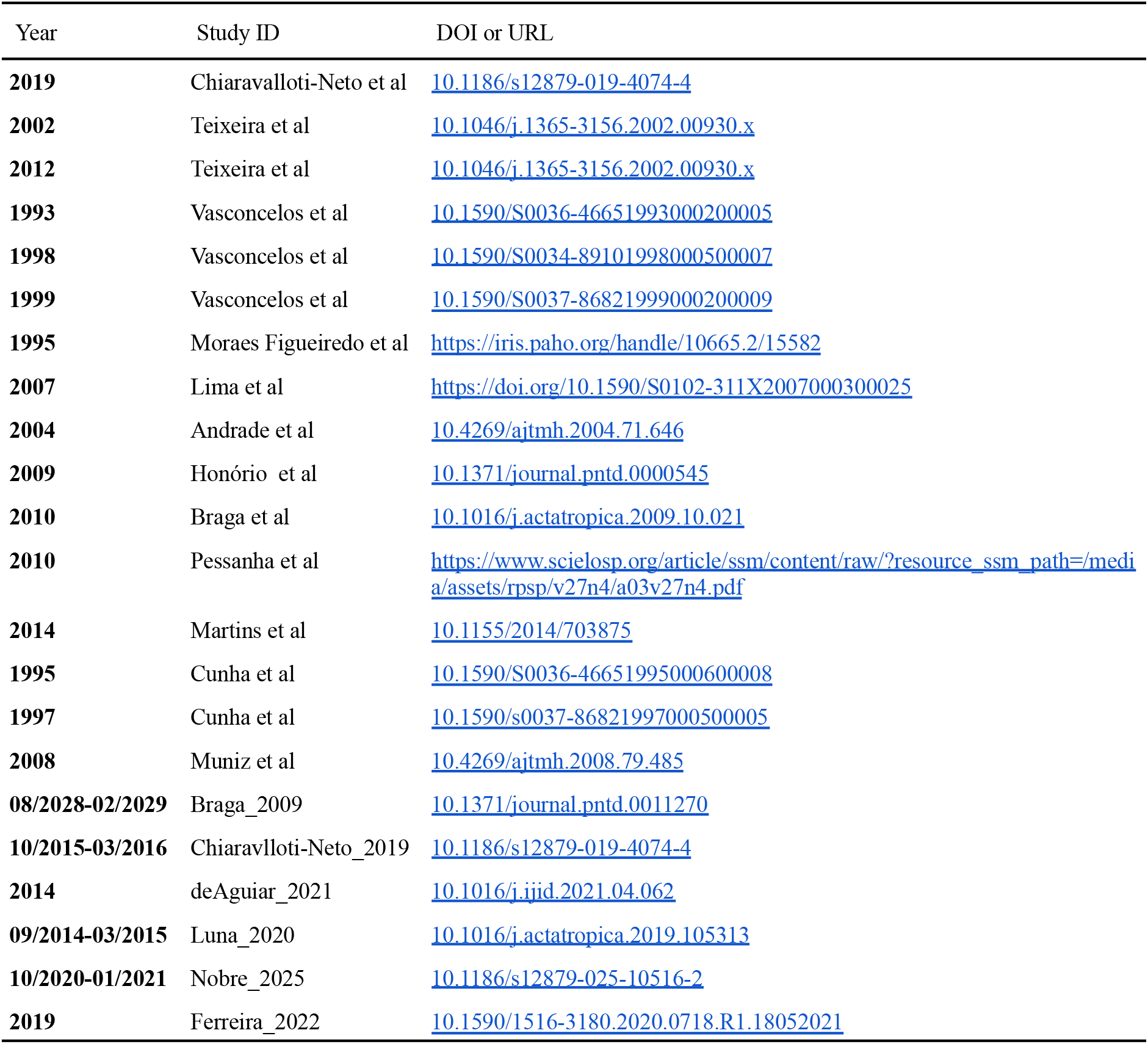
Seroprevalence studies used to fit the force of infection model.

**Figure S1.**
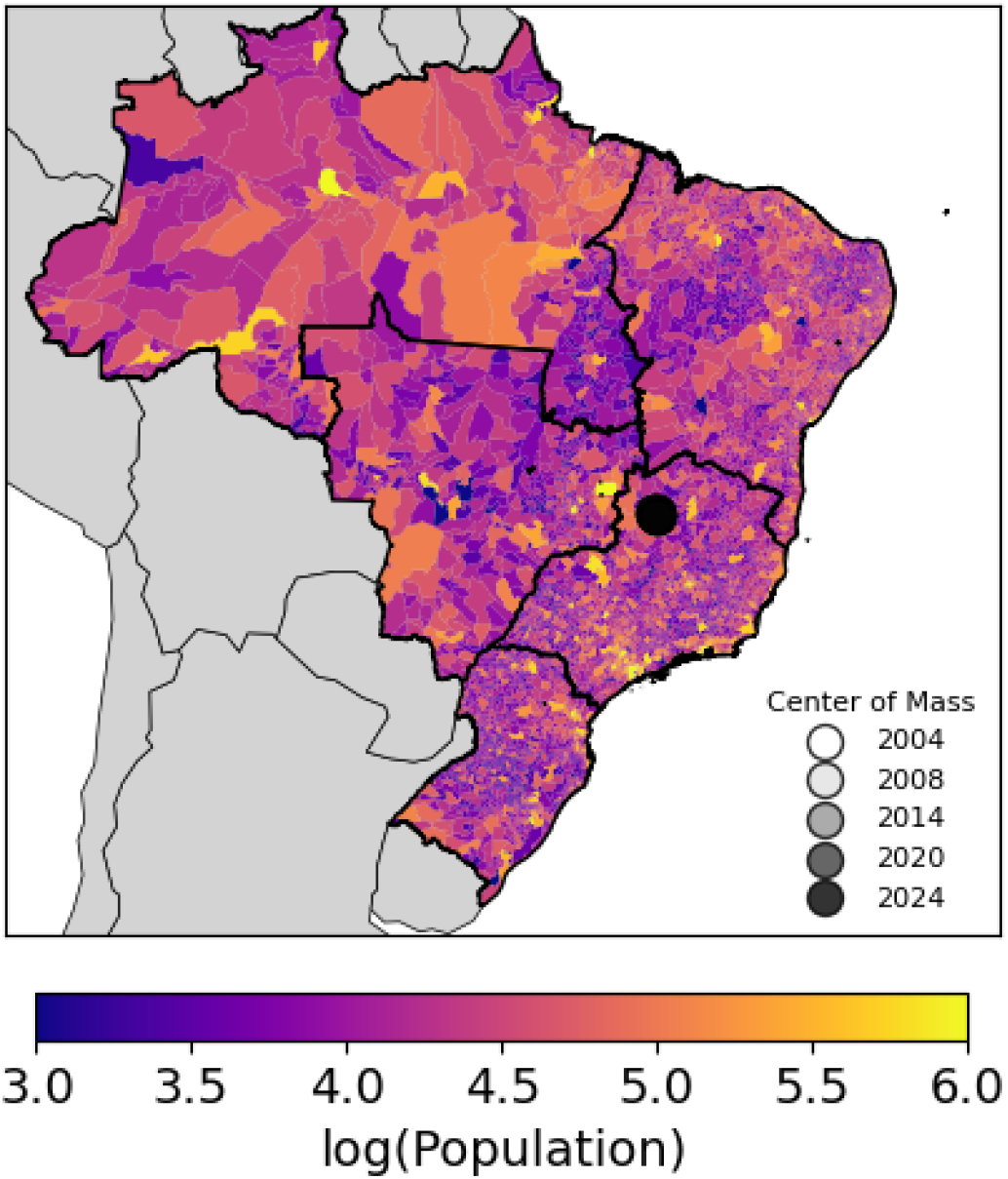
log of population obtained from DATASUS with the center of mass of population between 2004 and 2024. The center of mass does not change significantly with time.

**Figure S2.**
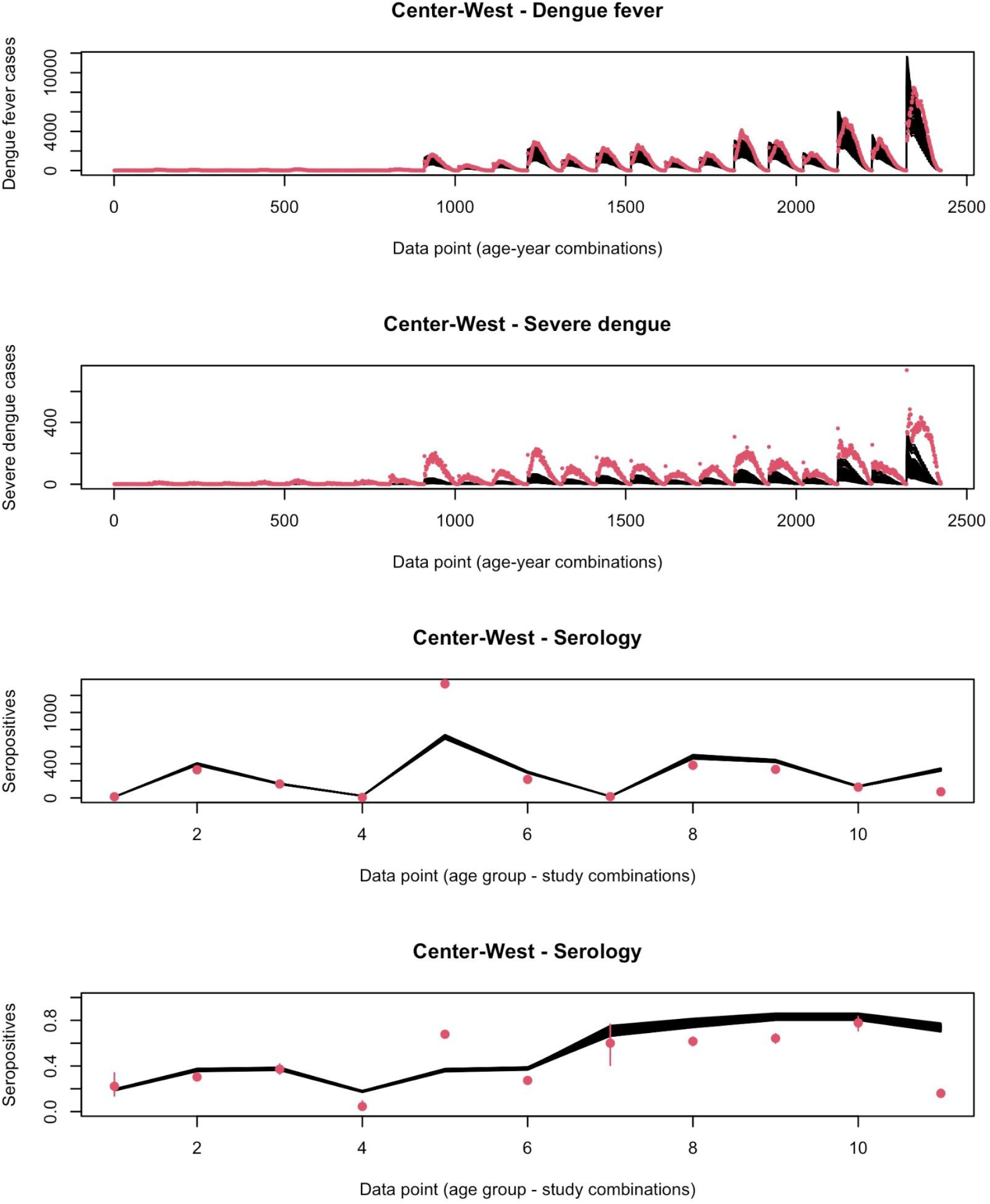
Comparison between the estimated number of dengue fever and severe dengue cases as well as serology by age group across the Centre-West region. The red dots represent real data and black lines realisations from the fitted model.

**Figure S3.**
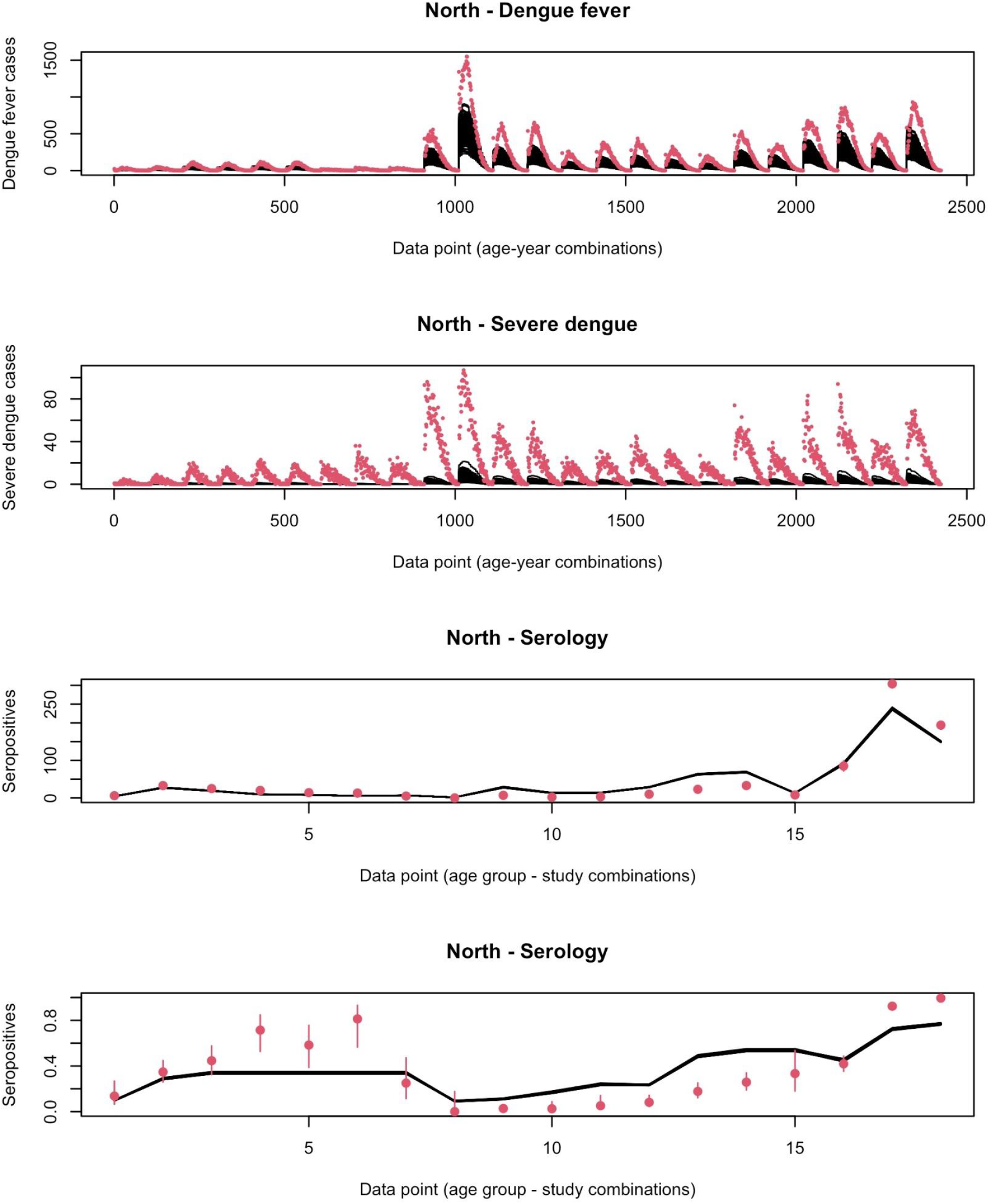
Same as Figure S2 but for the North region.

**Figure S4.**
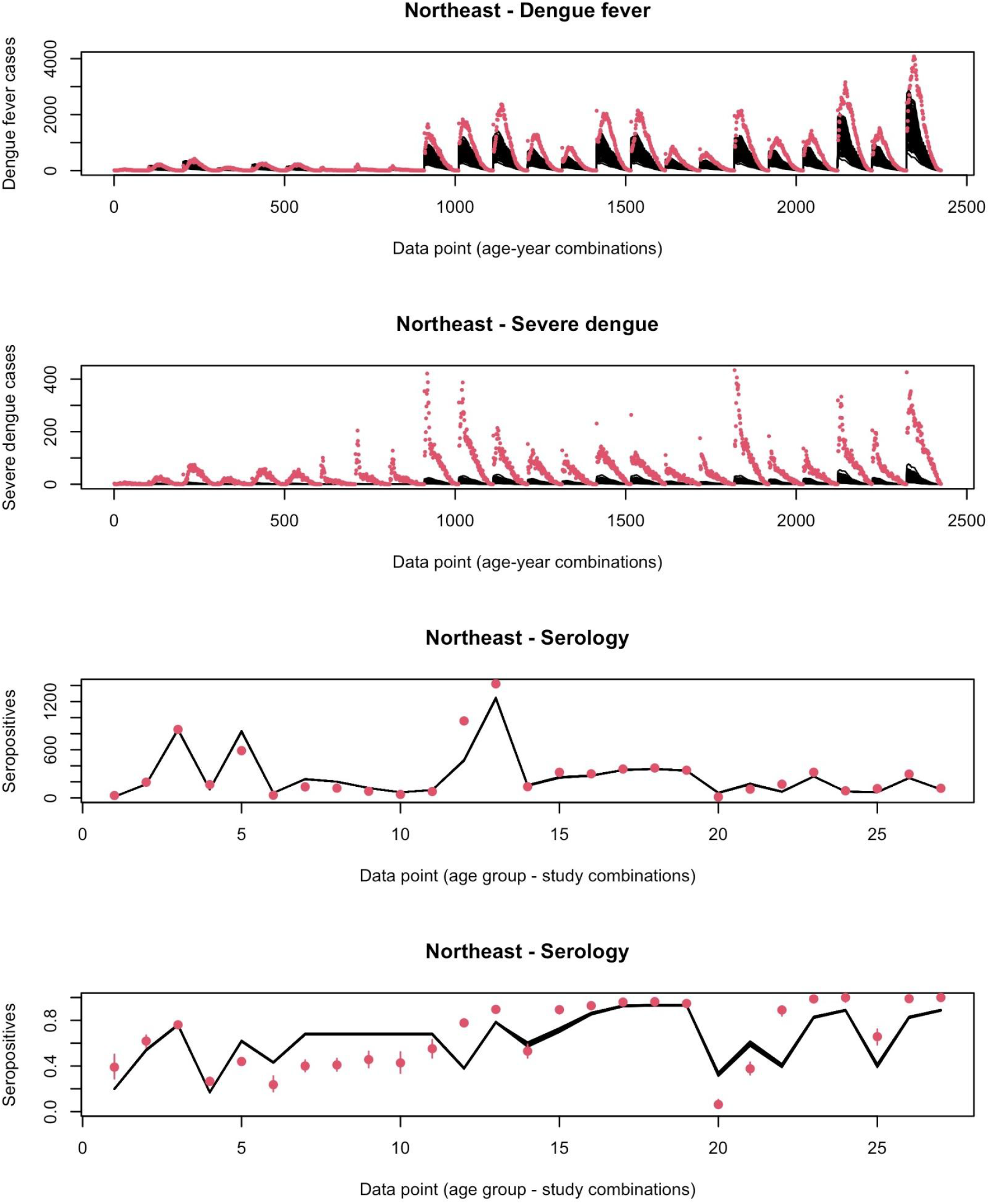
Same as Figure S2 but for the Northeast region.

**Figure S5.**
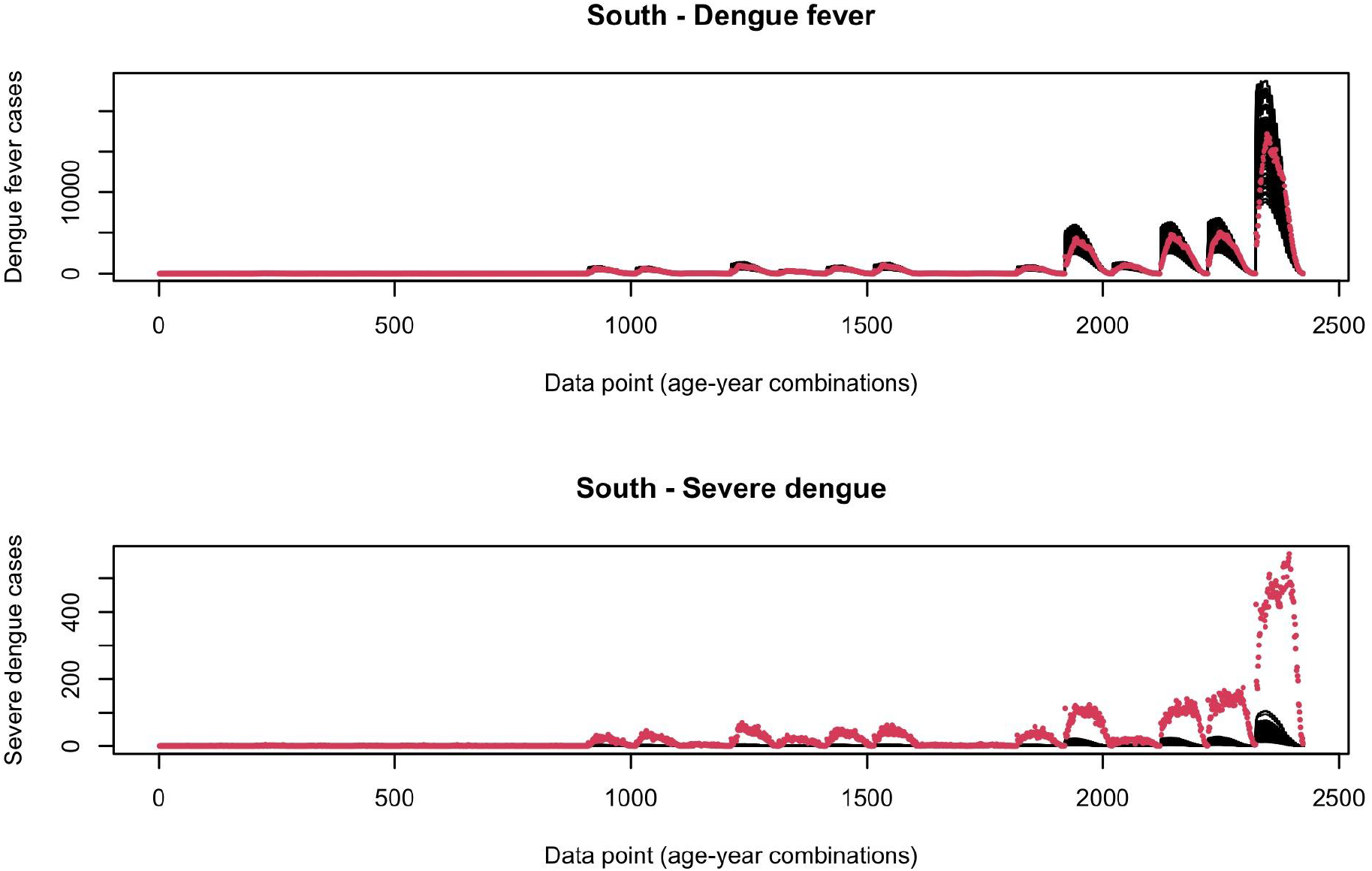
Same as Figure S2 but for the South region.

**Figure S6.**
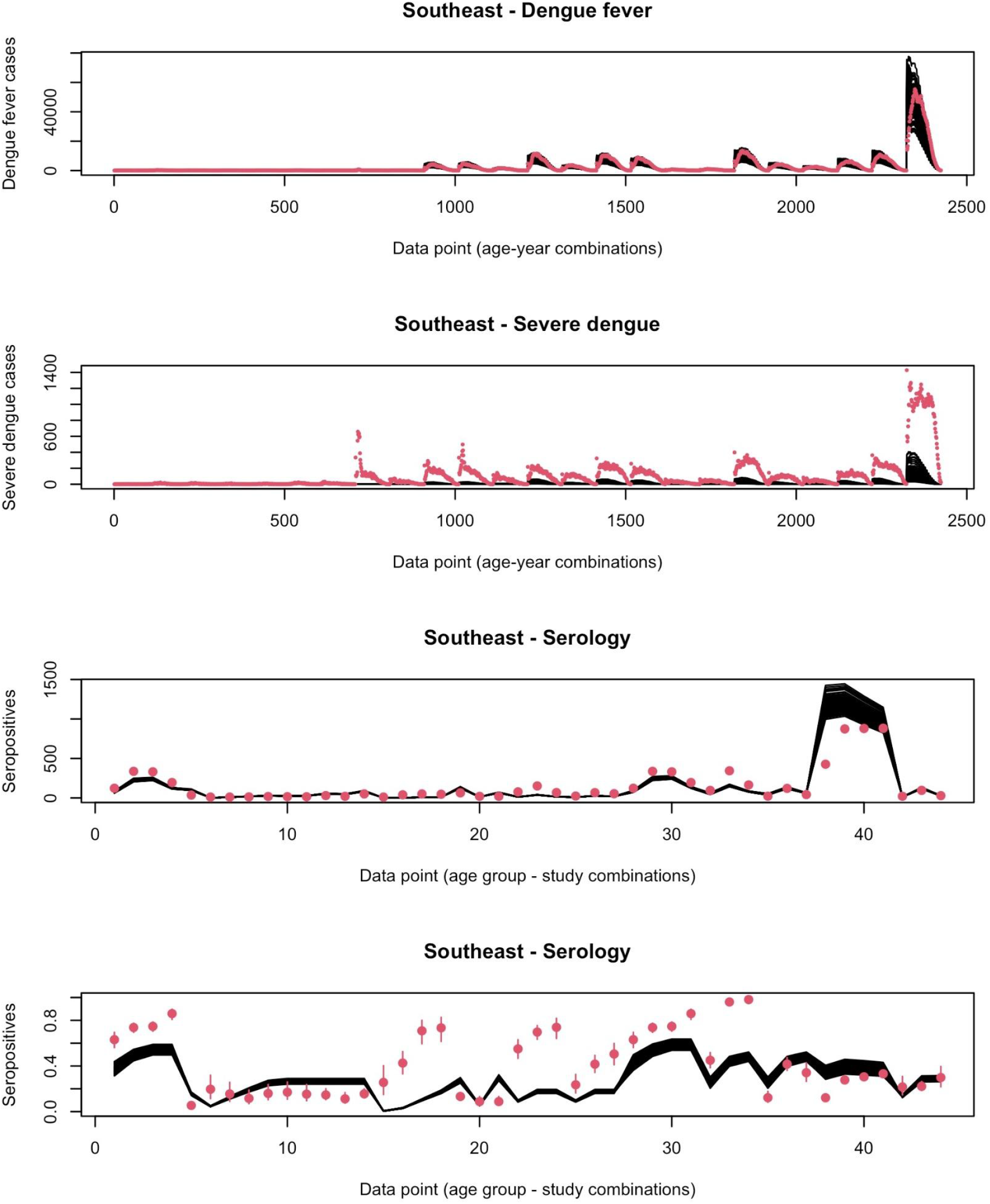
Same as Figure S2 but for the Southeast region.

**Figure S7.**
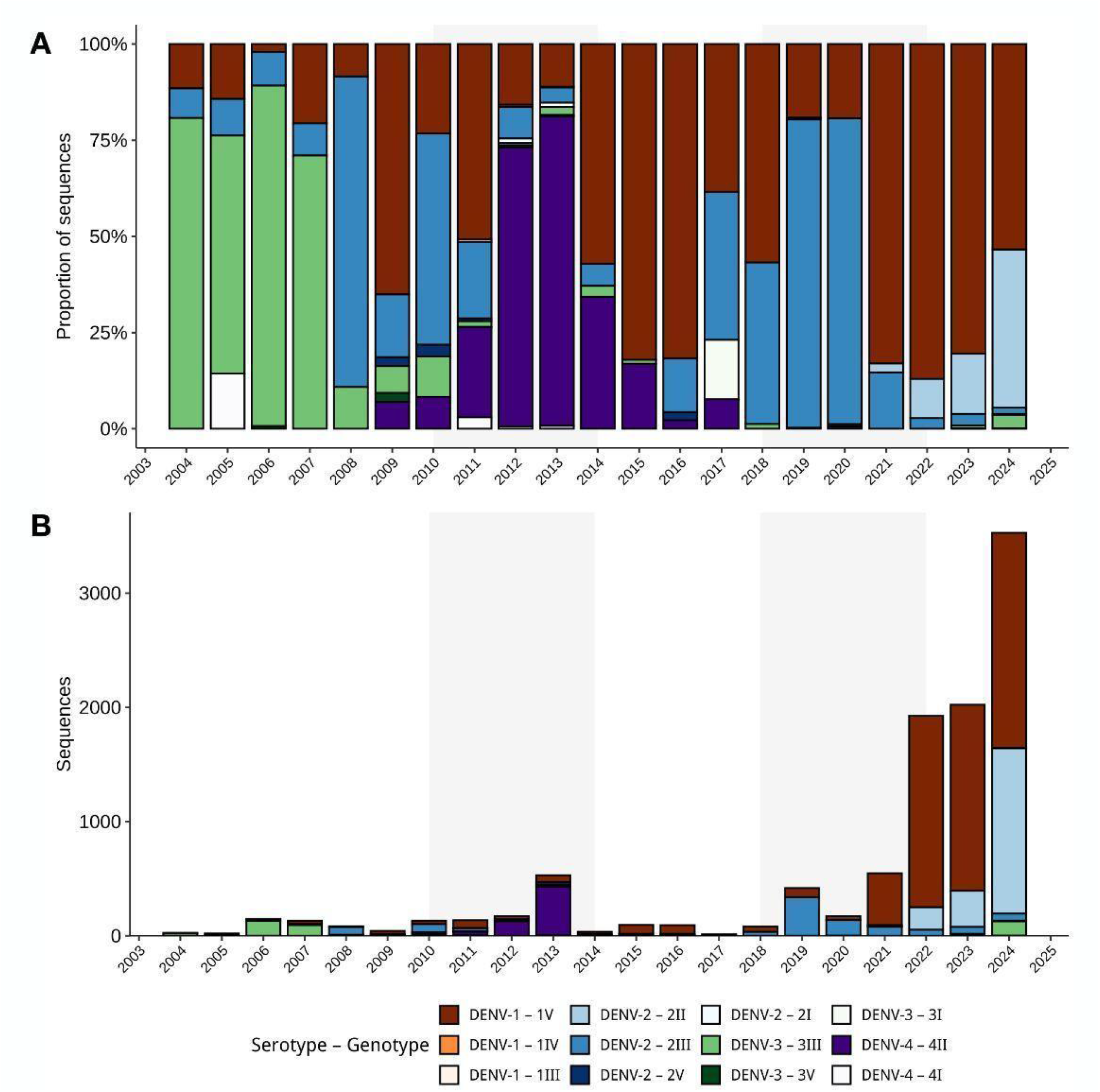
A) Annual genotype proportion of DENV in Brazil. B) Number of yearly DENV pathogen genomes by genotype in Brazil.

**Figure S8.**
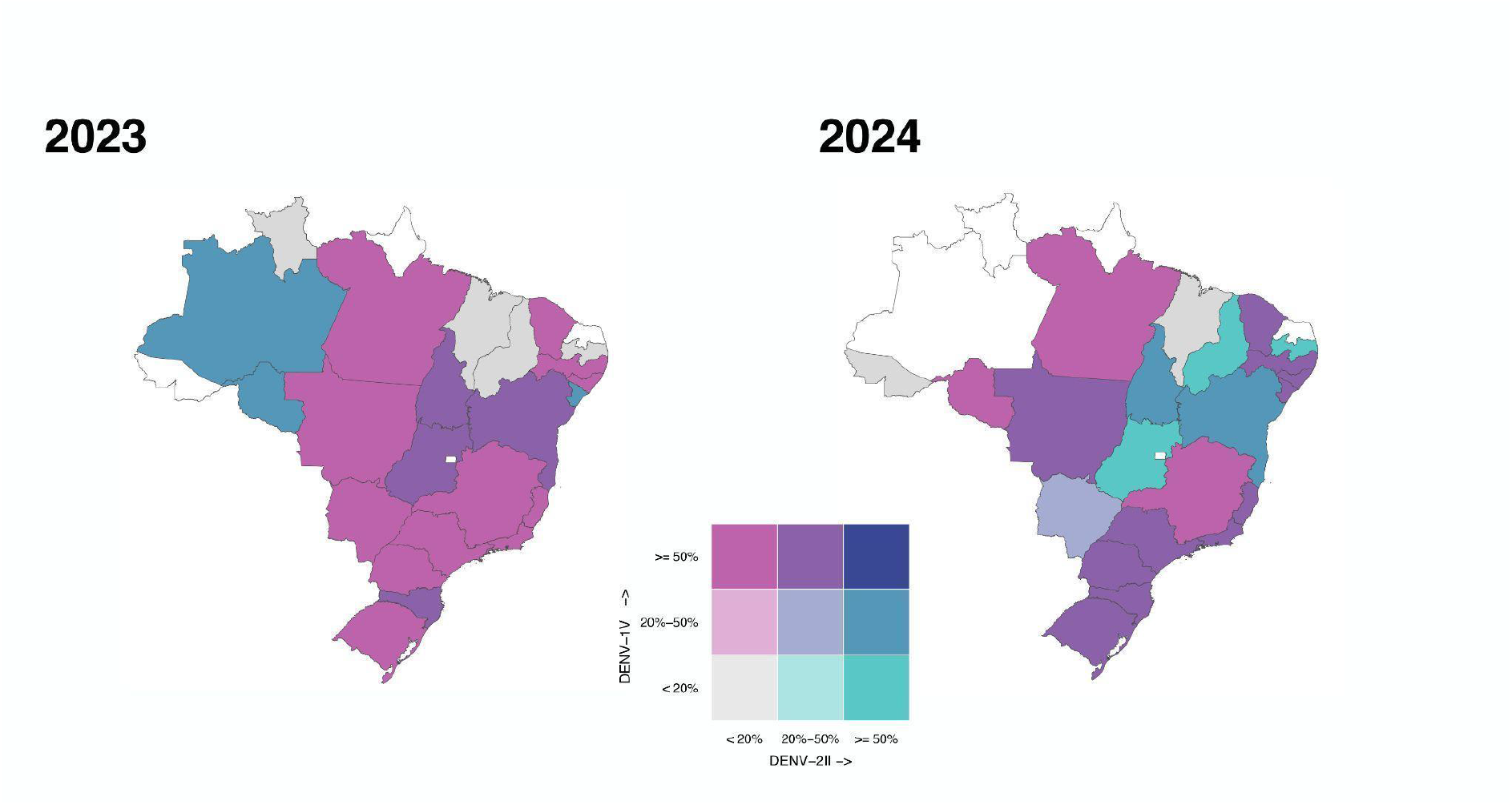
Bivariate choropleth map of Brazil at the state level showing the proportion of the two major circulating genotypes, DENV-1V and DENV-2II, in the years 2023 and 2024. The grey areas show states where less than five sequences were recorded, whilst the white areas are where there is no data available.

**Figure S9.**
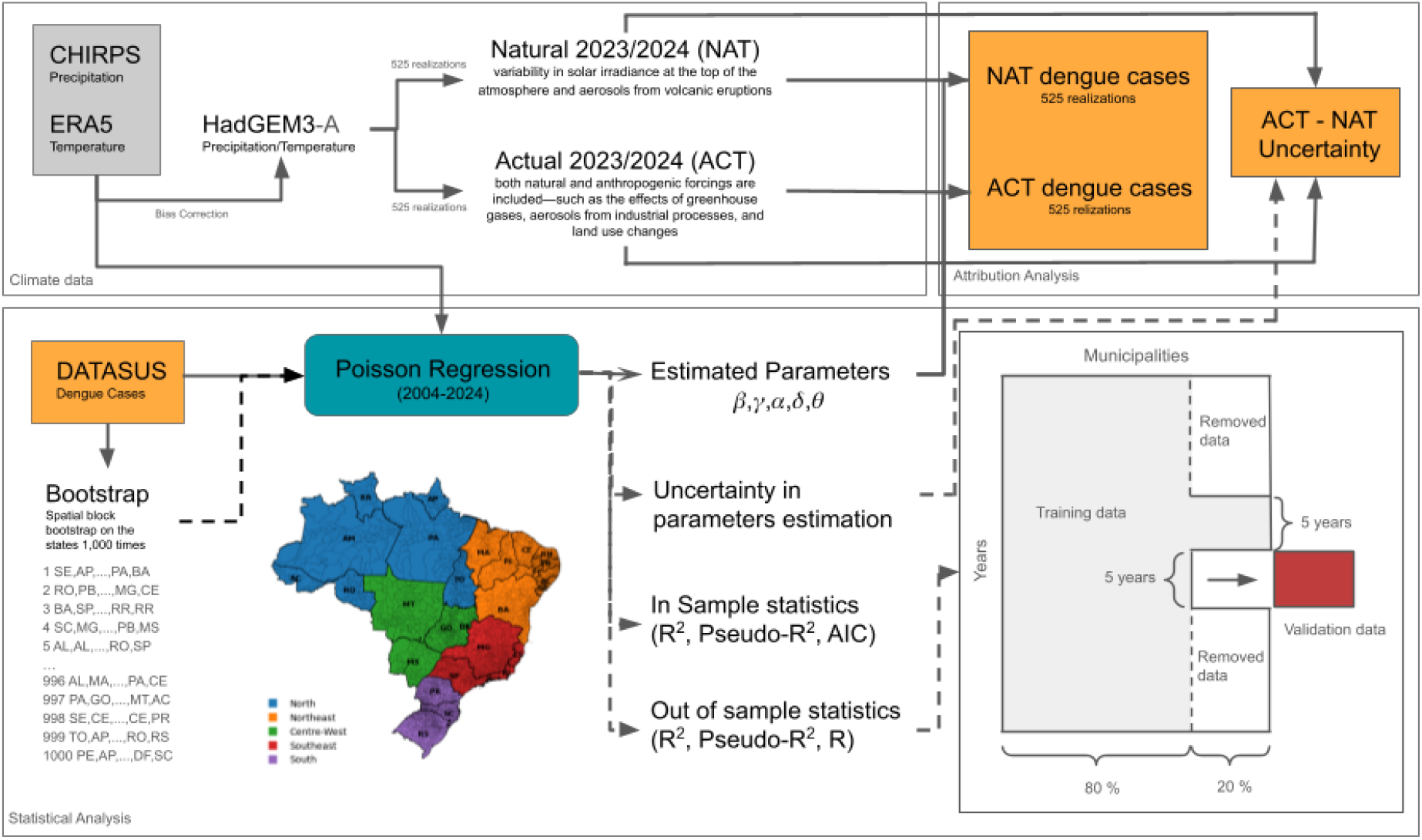
Flow chart of the pipeline for this study, with the datasets that were used, the statistical analysis, and attribution using ACT and NAT scenarios. We also add the uncertainty analysis that was performed based on spatial block bootstrap in dashed lines, and the out-of-sample validation diagram. For that we randomly select 20% of the municipalities to remove from the sample, just keeping 5 consecutive years that are randomly selected to calculate the fixed effects and estimate the parameters with the remaining 80 %. After that we validate the data on the following 5 years of the data for the 20 % of the municipalities that were removed.

**Figure S10.**
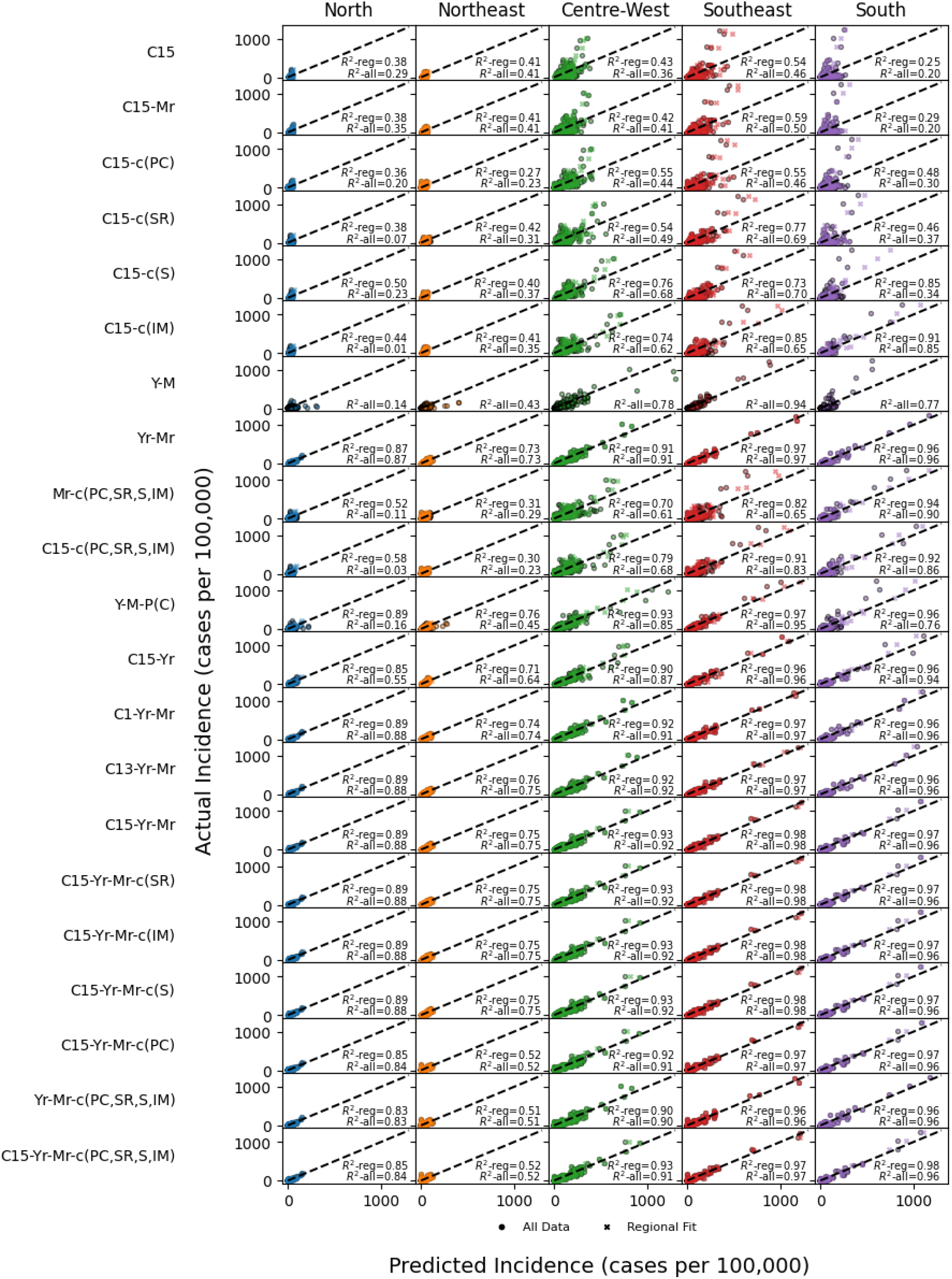
Scatter plot of predicted incidence versus actual dengue incidence aggregated for each region in Brazil for each of the tested models. The circle markers represent the fitted data using all observations, while the x markers represent the individual regional fits.

**Figure S11.**
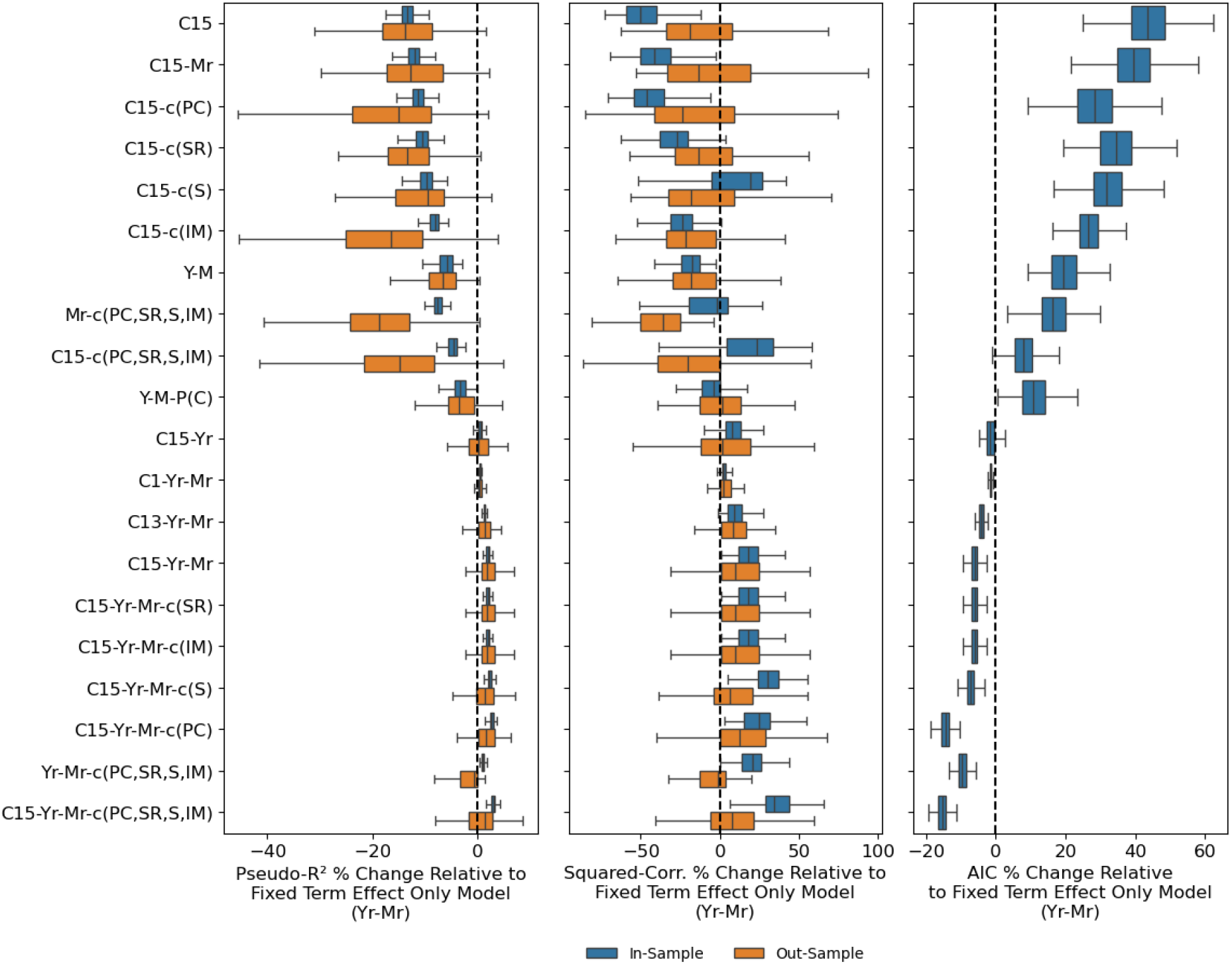
Change in Pseudo-R2, Squared Correlation and AIC for in-sample (blue) and out-sample estimates (orange) for the model with only fixed effect, i.e., only month, year, and municipality terms, without any climate variables, expressed as a percentage of change. The error bars are estimated from a spatial block bootstrap of 1,000 samples for in-sample and 100 samples for out-sample, sampling from the different states in Brazil;

**Figure S12.**
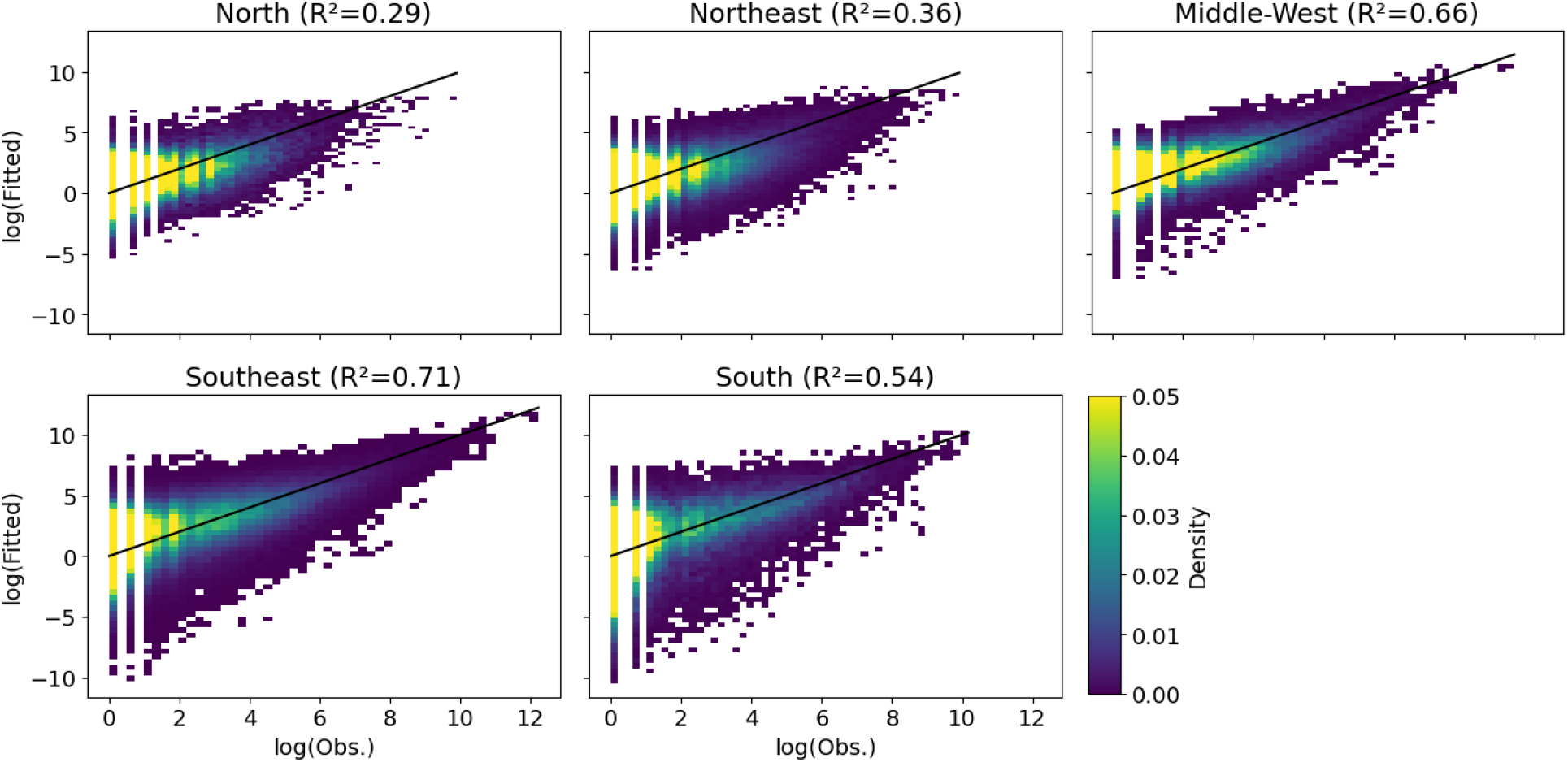
Scatter plot of the log of observed dengue incidence versus fitted dengue incidence (in sample). The scatter plot shows the point density representing each municipality in each month and year in the fitted data. The black line represents the Ordinary Least Squares (OLS) best fit and the R^2^ is between the number of observed cases and the fitted cases for each region.

**Figure S13.**
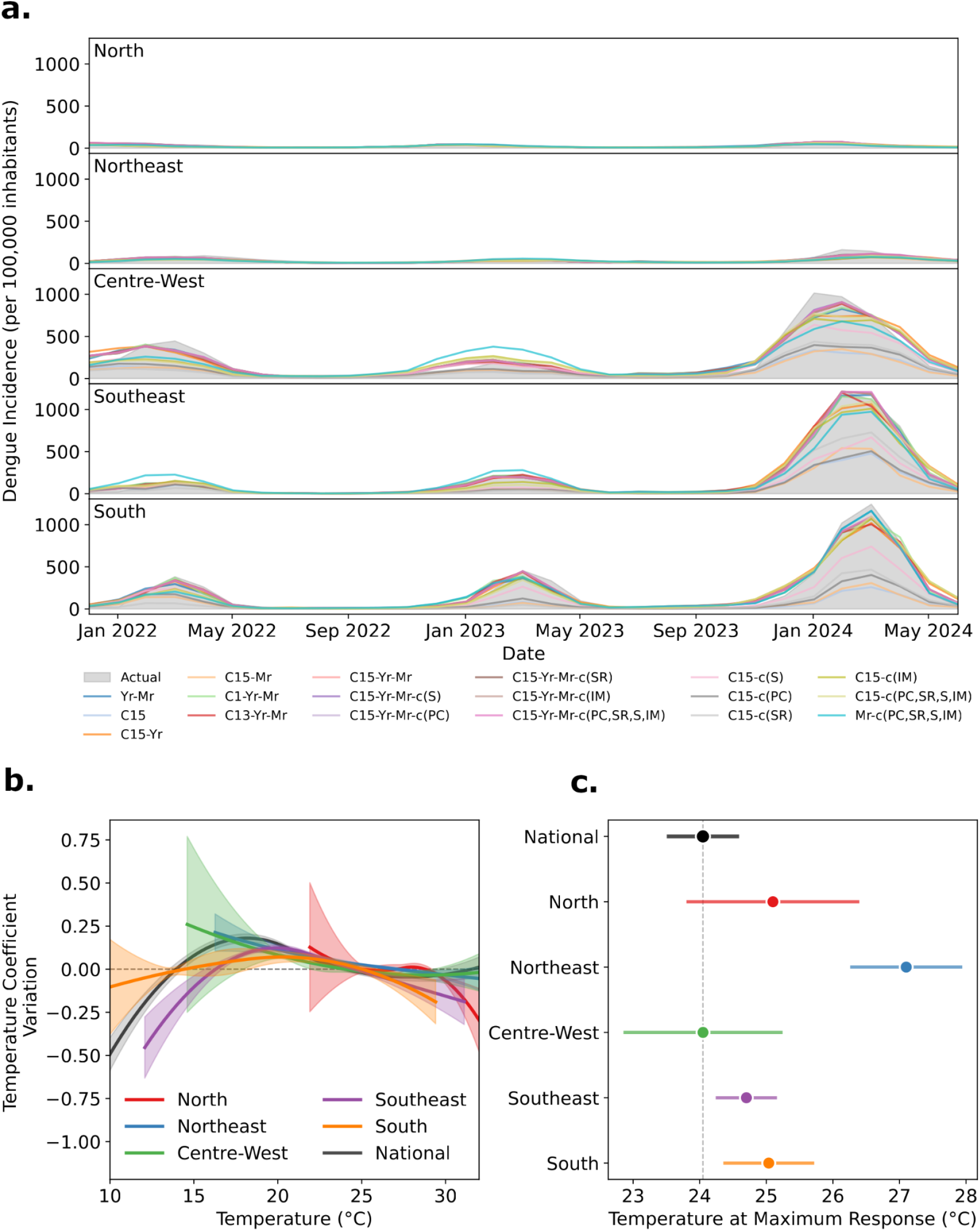
(a) Time series of monthly dengue incidence per 100,000 inhabitants for each region in Brazil. Grey shading represents the reported dengue incidence, while the different lines show the results of the model sensitivity tests fitted individually for each region; (b) Temperature–response curves for dengue, derived from the fitted Poisson regression model for each region using model C15-Yr-Mr-c(PC,SR,S,IM). Shading indicates the 95% confidence intervals calculated from analytic estimates; (c) Temperature corresponding to the maximum response for each regional fit shown in (b). The 95% confidence intervals are also derived from analytic estimates.

**Figure S14.**
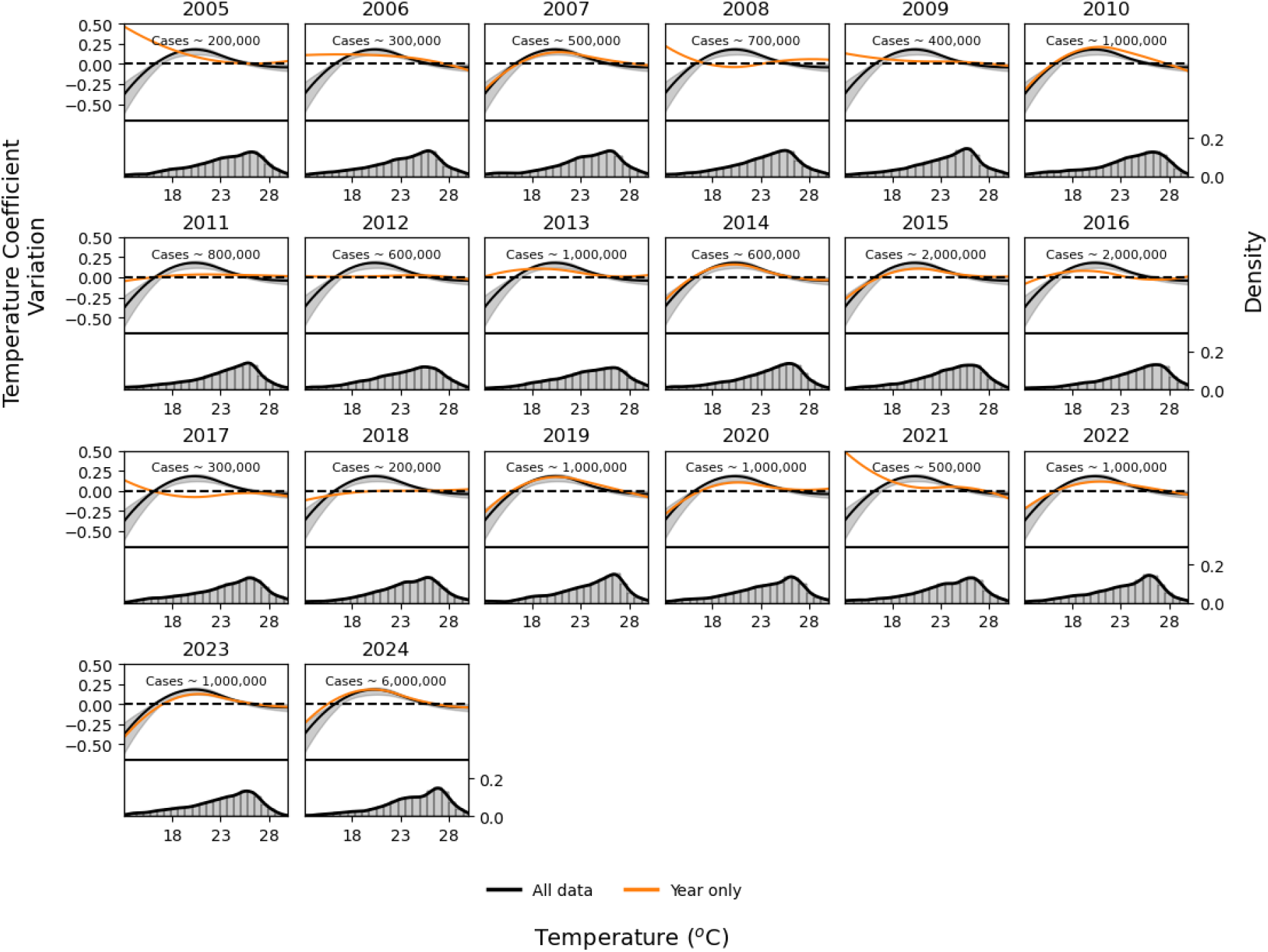
Temperature response in the Poisson regression model for each year (orange) and for all years (black line). The shading shows the 95% confidence intervals calculated using 1,000 bootstrap samples. In the bottom of each plot is the temperature histogram for each year. At the top, the approximate number of cases for each year is shown.

**Figure S15.**
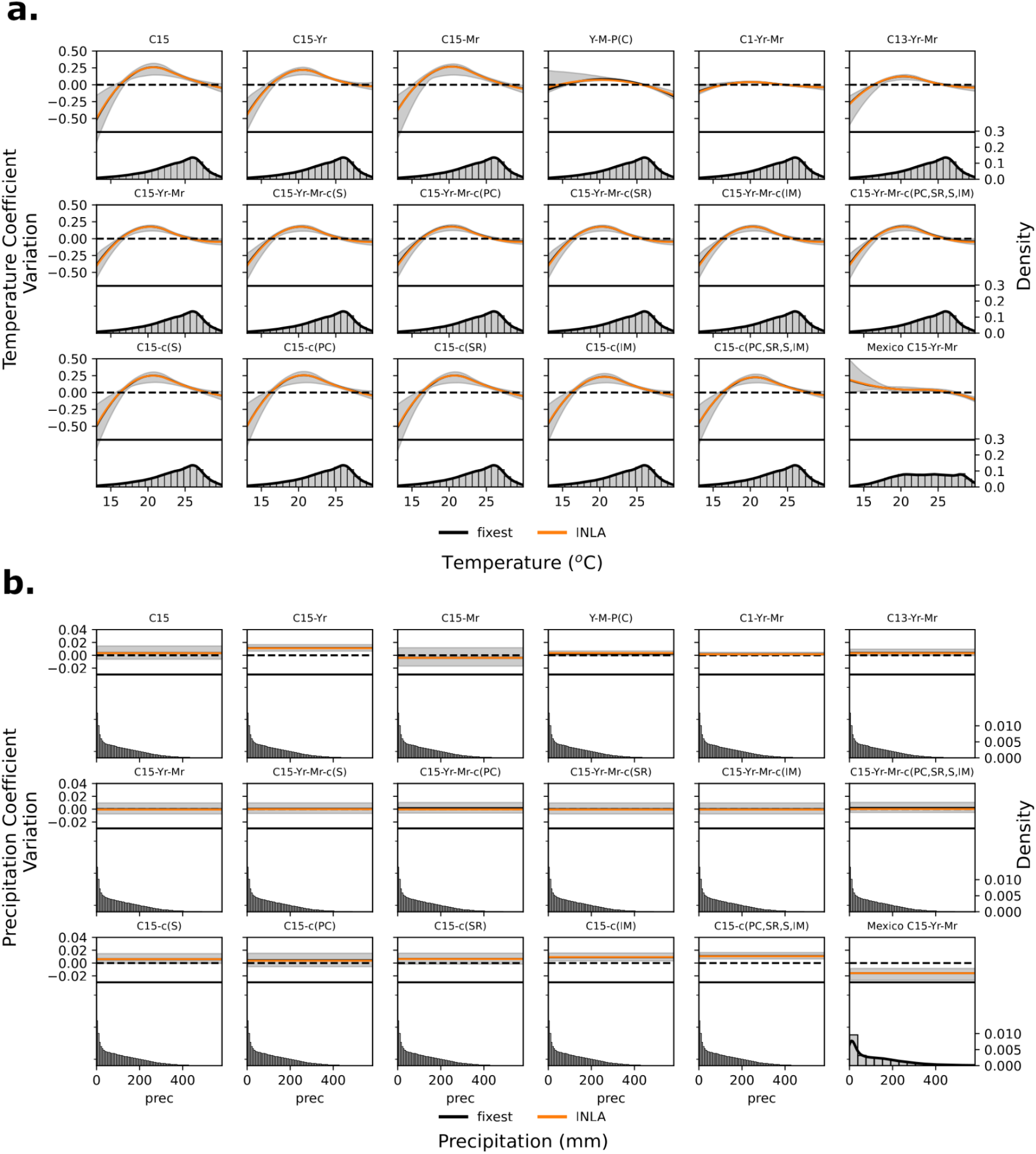
(a) Temperature response in the Poisson regression model shown through the coefficient variation with temperature. The shading shows the 95 % confidence intervals calculated using 1,000 bootstrap samples. Black line represents the model fitted using fixest package and in orange the INLA model with random effects assumed to be iid variables for municipality and year, and a cyclic random walk term for the month ; (b) Same as (a) but for precipitation coefficients.

**Figure S16.**
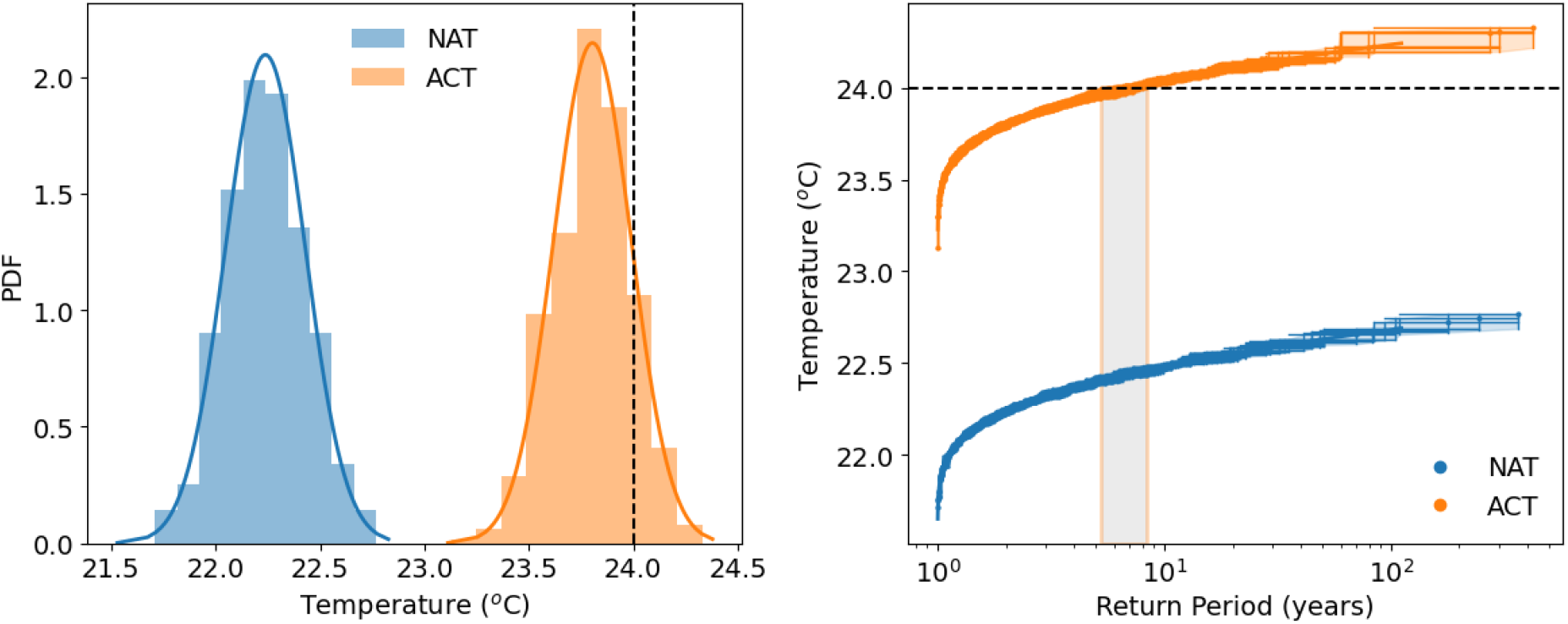
(left) Histogram of population weighted temperature in Brazil for the 525 ensembles of ACT and NAT scenarios. The line represents a fitted normal distribution. The dashed vertical line represents the observed value according to the ERA5 dataset; (right) Return period of population weighted temperature for ACT and NAT scenarios. The dashed horizontal line is the observed value according to the ERA5 dataset.

**Figure S17.**
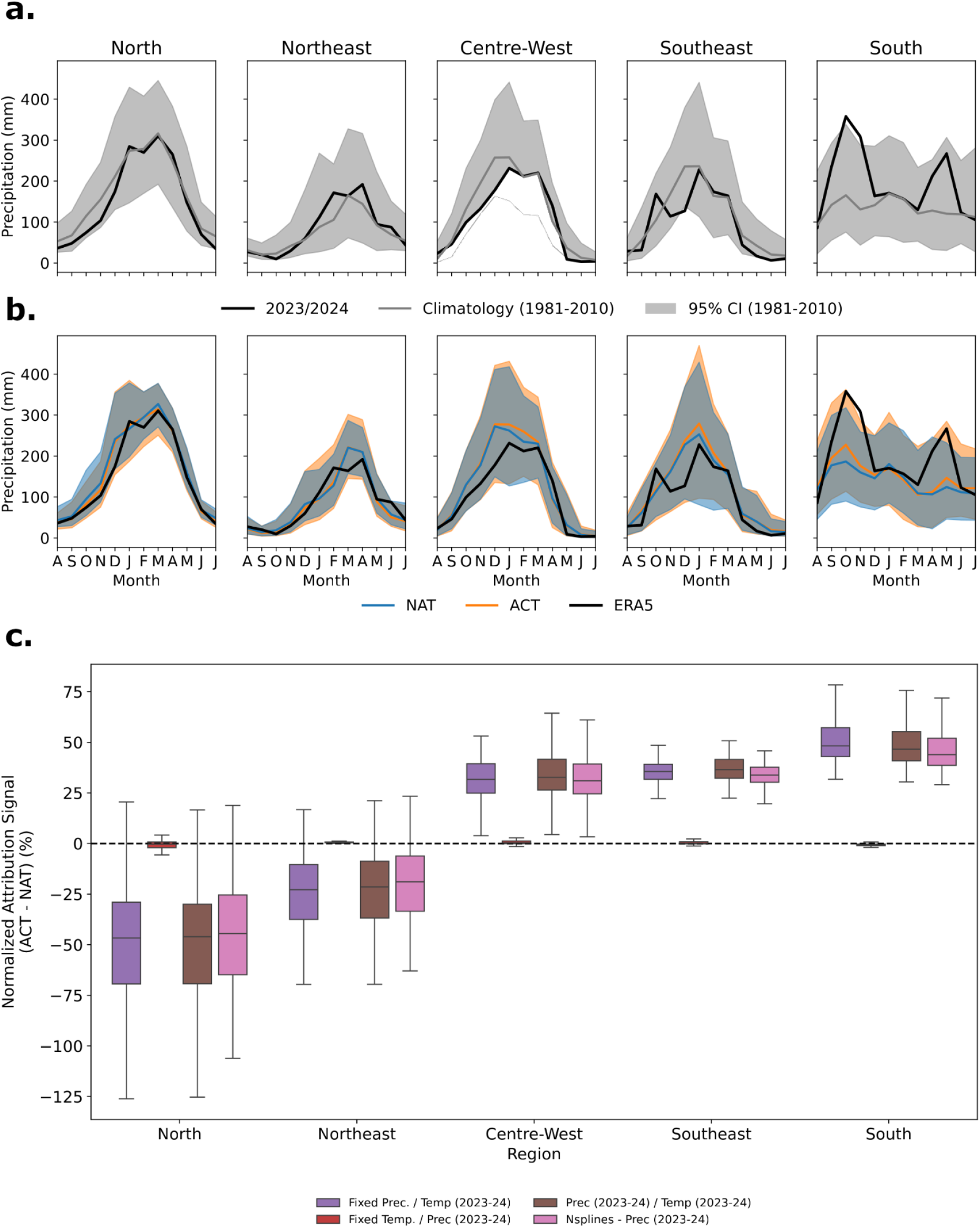
(a) Mean CHIRPS precipitation for each region between August 2023 and July 2024, compared to the climatology calculated for the period 1981–2010; (b) Comparison of observed precipitation from CHIRPS in 2023/24 with respect to model simulations from HadGEM3-A under scenarios with anthropogenic climate change (ACT) and without (NAT); (c) Percentage differences in estimated dengue cases using: fixed temperature based on the monthly mean for 2002–2015 with precipitation set to 2023/24 (red bar); fixed precipitation with temperature set to 2023/24 (purple bar); both temperature and precipitation set to 2023/24 (brown bar); and using natural splines instead of a linear fit to model precipitation (pink bar).

**Figure S18.**
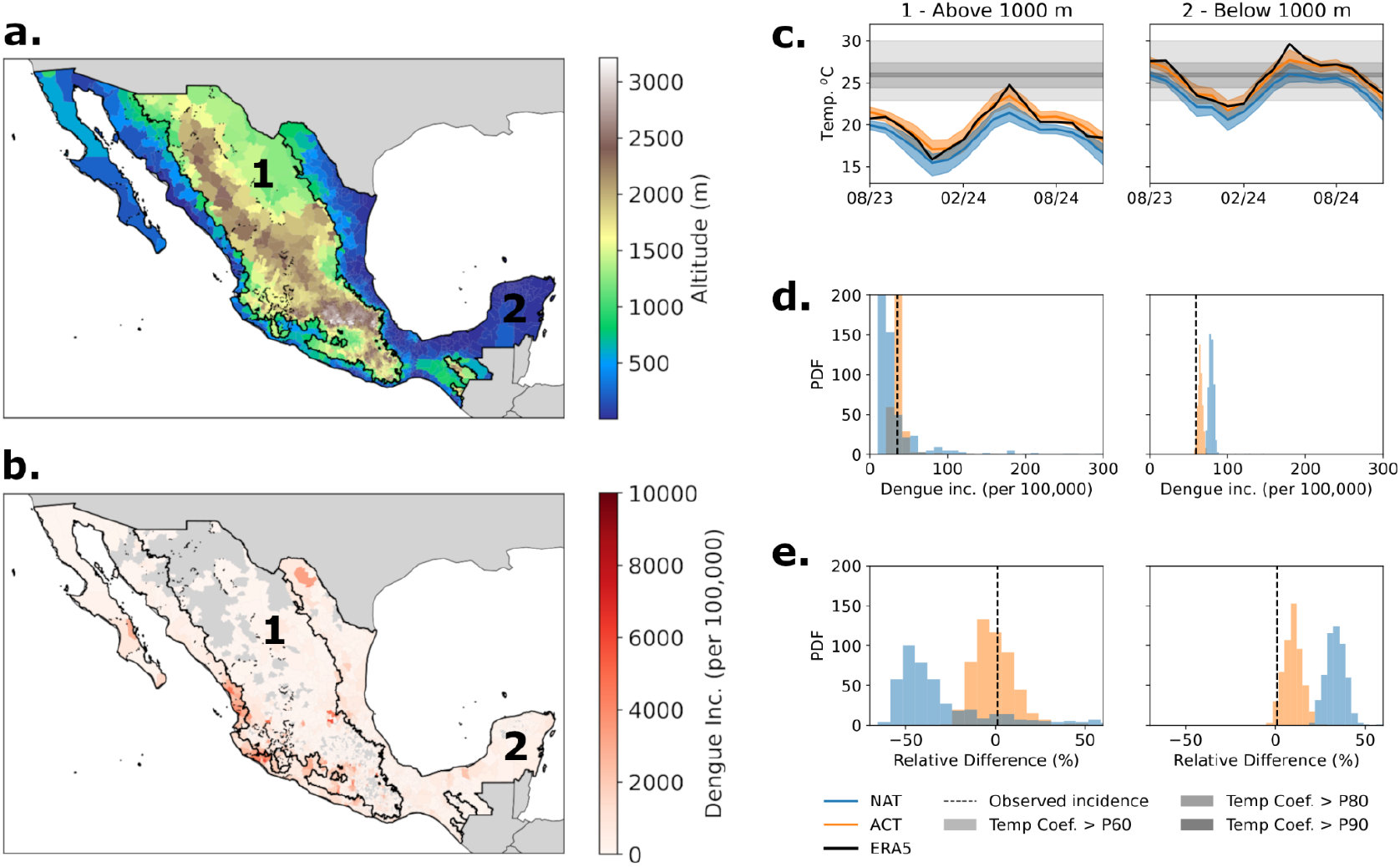
(a) Map of altitude in Mexico; (b) Map of the dengue incidence (per 100,000 inhabitants) between January and December 2024 for Mexico; (c) Average temperature in low and high altitude regions in Mexico (1 being high and 2 being low), for the ACT and NAT simulations from HadGEM3-A model compared with ERA5. The shadings show the ranges of estimated thermal optima for dengue transmission based on the temperature response from the fitted model using data from Mexico and Brazil; (d) Histogram of the dengue incidence for each one of the 525 ensemble members in the ALL and NAT simulations in cases per 100,000 inhabitants; (e) Same as (d) but as the difference between ACT (NAT) and the observed number of cases, divided by the observed incidence.

**Figure S19.**
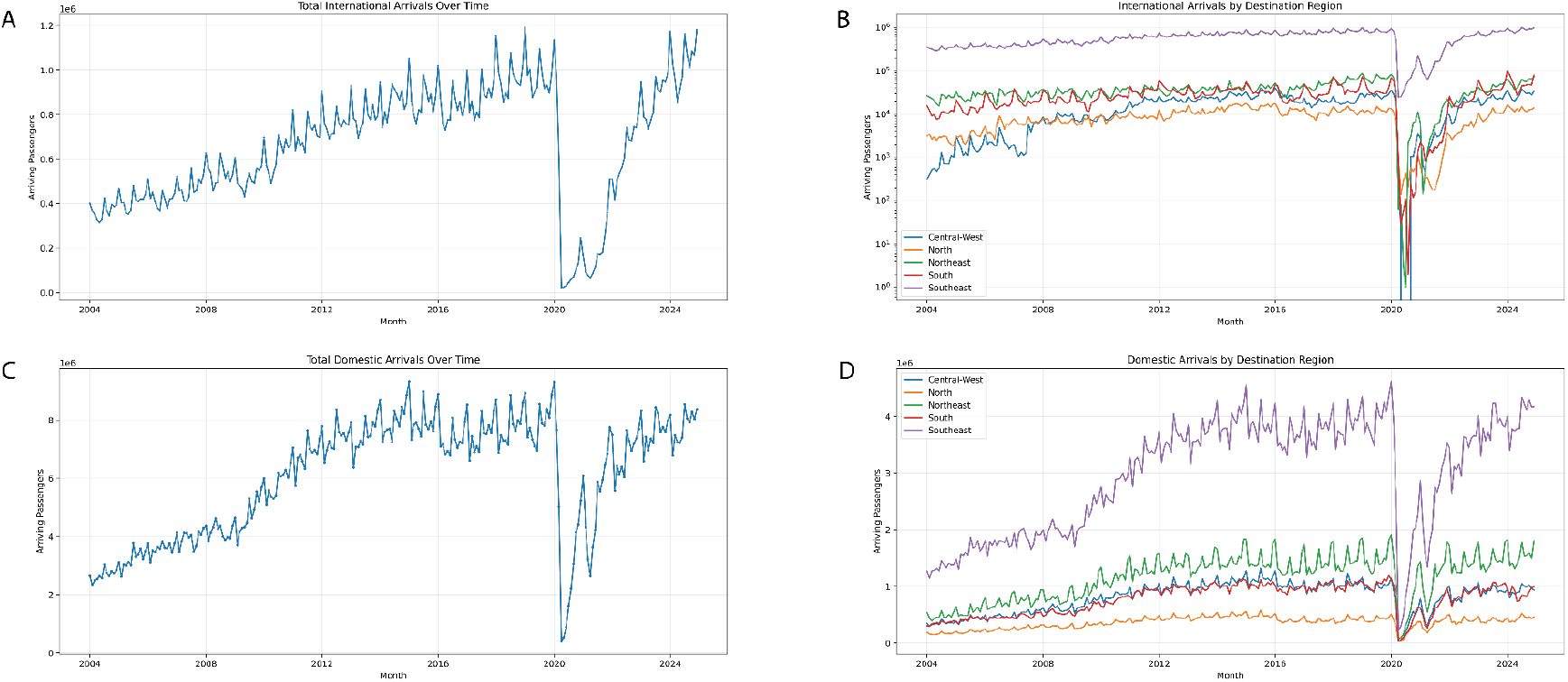
Summary of international arriving passengers in Brazilian airports from the Agência Nacional de Aviacao Civil (Agência Nacional de Aviação Civil (Anac), n.d.). (a) International arrivals of passengers on direct flights into airports in Brazil. (b) Disaggregated by region in Brazil. (c) Domestic arriving passengers in Brazil. (c) Disaggregated arriving passengers by region in Brazil.

**Figure S20.**
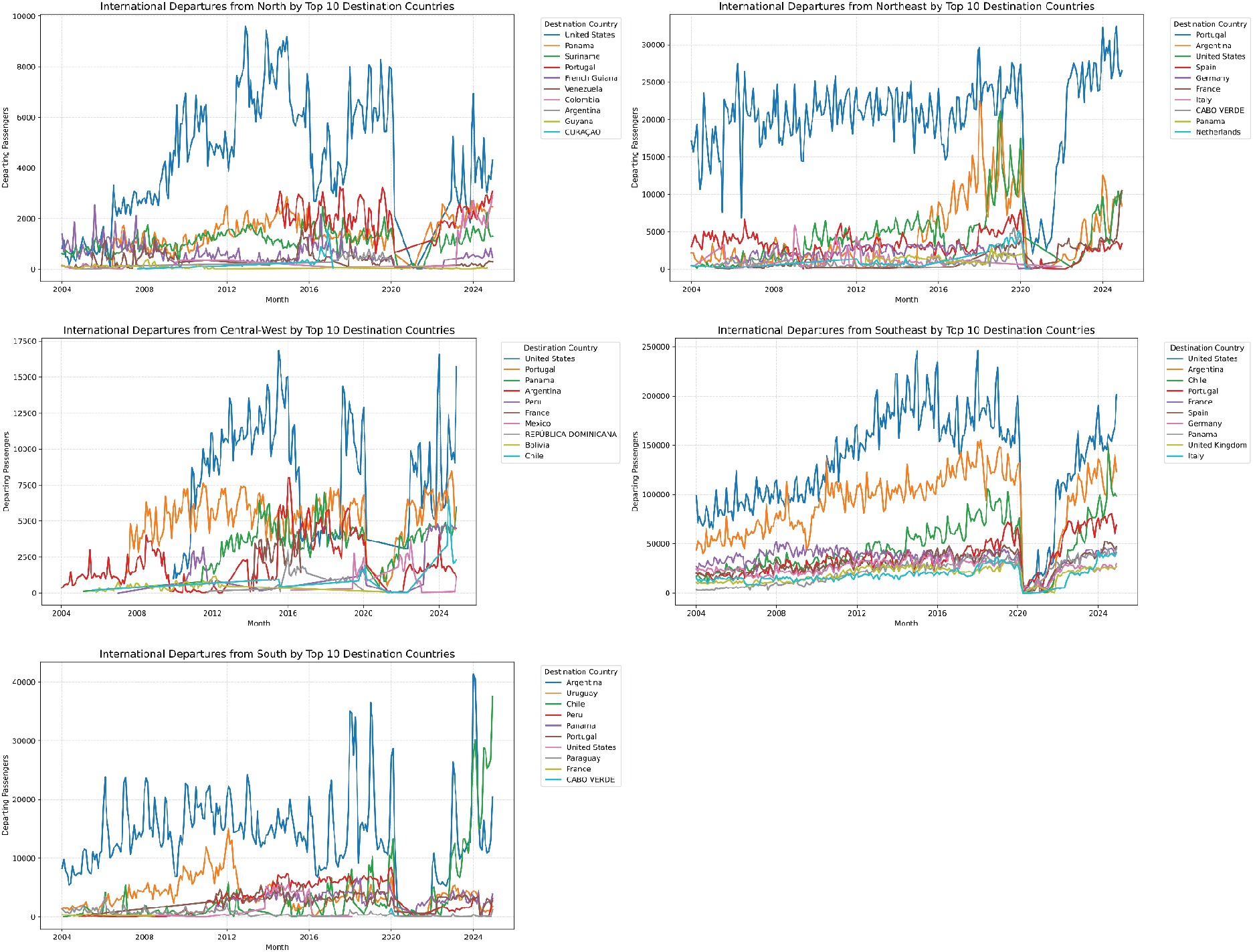
Summary of departing passengers from Brazilian airports to the top 10 countries per region. Data from (Agência Nacional de Aviação Civil (Anac), n.d.).

**Figure S21.**
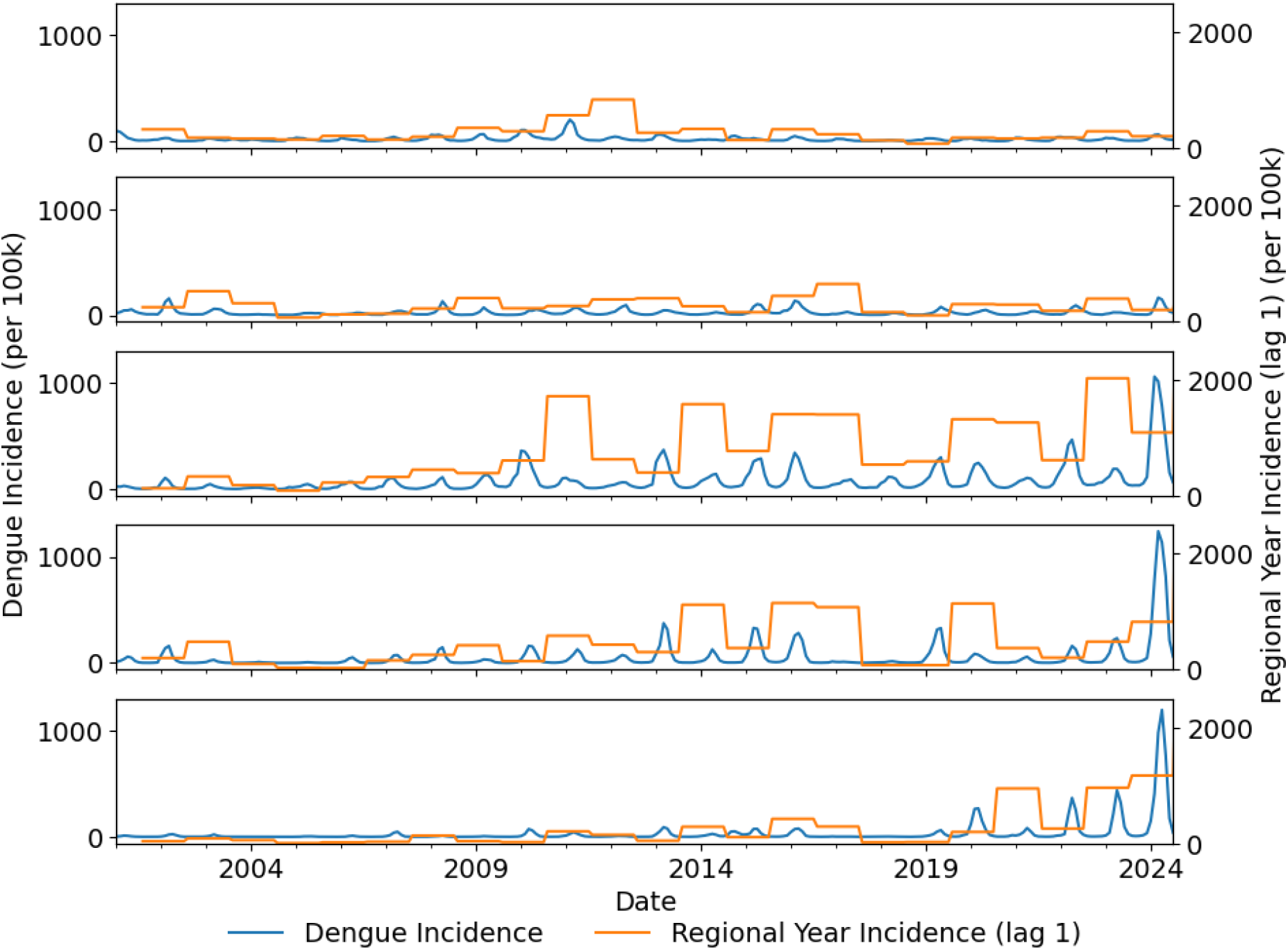
Monthly dengue incidence per 100,000 inhabitants (blue line) and the aggregated dengue incidence for each region lagged one year to serve as an immunity proxy.

**Figure S22.**
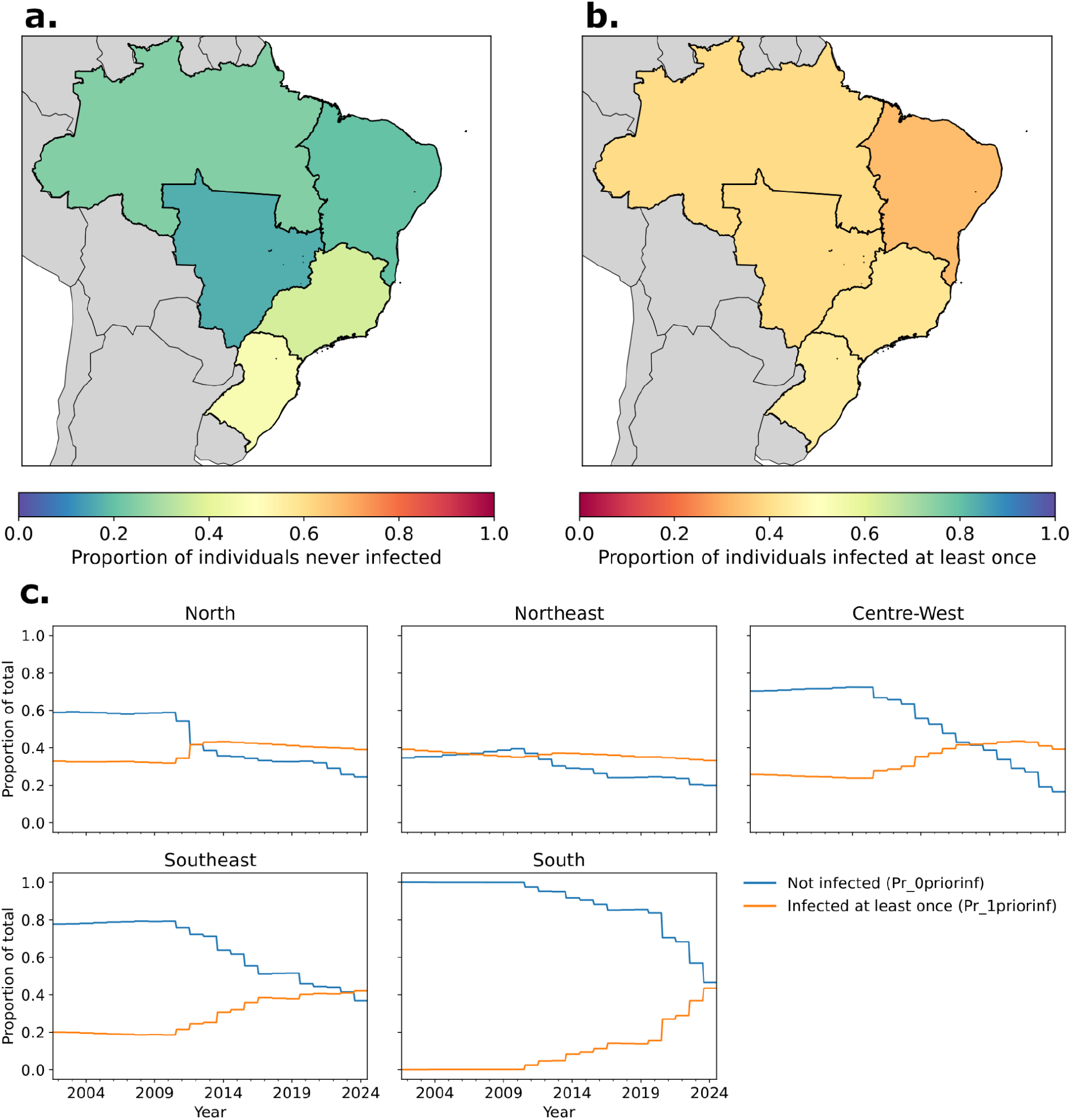
Proportion of the population never infected (a) and infected at least once (b) ahead of the 2023/24 season and annually (c).

